# Modeling pathway overlap increases accuracy of GWAS gene set enrichment

**DOI:** 10.64898/2026.09.03.26362206

**Authors:** Alanna C. Cote, Reagan Kesting, Judit García-González, Paul F. O’Reilly

## Abstract

Genome-wide association studies (GWAS) have identified thousands of loci associated with complex traits and diseases, and extensive efforts are underway to translate these variant-level signals into biological mechanism. A widely applied approach is pathway enrichment analysis, which tests whether genetic associations concentrate within biological pathways beyond background polygenic expectations. However, pathway databases contain extensive sharing of genes across pathways (“pathway overlap”), an underappreciated source of bias that creates structural dependencies in enrichment statistics and obscures pathway-specific genetic signal. Moreover, the degree of pathway overlap is increasing as pathway resources expand. Here, we introduce Gene Swap Randomization (GSR), an empirical framework that preserves pathway size and multi-pathway gene membership in the null model, enabling explicit adjustment for pathway overlap. Applying GSR to enrichment results from the Molecular Signatures Database (MSigDB) across twelve complex traits and four pathway analysis approaches (MAGMA, PascalX, GSA-MiXeR, and PRSet), we show that pathway overlap can produce enrichment under polygenicity even in the absence of pathway-specific biology. GSR improves prioritization of biologically relevant pathways supported by independent gene-disease associations (Open Targets, Malacards), regulatory interactions (DoRothEA), and tissue-specific expression patterns (GTEx). GSR improves concordance with external benchmarks in 60.8% of comparisons overall and 79.3% disease-association benchmarks, corresponding to improvement in 10 of 16 aggregated method-validation framework comparisons. We demonstrate that pathway overlap is a key source of bias in GWAS pathway enrichment, that pathway-specific disease enrichment persists after conditioning on overlap, and that GSR improves biological insight by distinguishing pathway-specific genetic signal from enrichment driven by pathway overlap.

## Introduction

Gene set or (hereafter) pathway enrichment analysis has become central to interpreting GWAS, enabling the transition from associated loci to biological mechanisms. These approaches test whether genetic associations concentrate within groups of functionally related genes, such as biological pathways or regulatory modules, beyond background polygenic expectations. GWAS pathway enrichment analyses typically use a competitive framework that tests whether genes within a pathway are more strongly associated with the phenotype than genes outside the pathway^1,2^, to evaluate whether genetic signal is concentrated within specific pathways relative to the genomic background.

A key but underexplored challenge of this framework is pathway overlap, in which genes are members of multiple pathways. This overlap may arise due to database redundancy, literature bias, or genuine biological multifunctionality. Importantly, pathway overlap has increased over time as pathway resources have expanded^3^. Despite this, many GWAS pathway enrichment methods estimate statistical significance using gene randomization^4,5^, sample randomization^6–8^, or regression-based frameworks^9^, which do not account for gene sharing among pathways.

Pathway overlap introduces several challenges for pathway enrichment analyses. Firstly, it creates statistical dependence among pathway tests, complicating multiple testing correction. Secondly, it generates clusters of closely related pathways that may all be flagged as biologically interesting, requiring *post hoc* pruning or conditioning on overlapping pathways to prioritize findings^10–12^. Lastly, pathway overlap may bias enrichment statistics, leading to either inflation or deflation of results and misleading interpretation of the importance of pathways to complex trait etiology. Prior work in transcriptomics has shown that downweighting or iteratively removing multi-pathway genes can improve the accuracy and reliability of pathway enrichment results^13,14^, but the impact of pathway overlap on GWAS enrichment remains poorly characterized.

In this study, we systematically quantify pathway overlap across a major pathway resource (MSigDb) and evaluate its impact on GWAS pathway enrichment results. We introduce Gene Swap Randomization (GSR), a novel framework for producing empirical null distributions of pathway enrichment tests by generating randomized pathways while preserving pathway size, gene membership frequency, and the global structure of pathway overlap (see Methods). Applying GSR, we evaluate whether adjustment for pathway overlap improves the accuracy of pathway enrichment results across twelve traits and four pathway analysis approaches (**Fig 1**). We show that GSR effectively adjusts enrichment statistics, re-ranking pathways and increasing their concordance with external benchmarks, including functional annotations and gene-disease evidence. We provide a user-friendly GSR tool, which only requires GWAS summary statistics and pre-defined pathways, enabling researchers to easily incorporate adjustment for multi-pathway genes into any GWAS pathway enrichment workflow, broadly supporting the accurate identification of biologically meaningful pathways underlying genetic association results.

**Figure 1.**
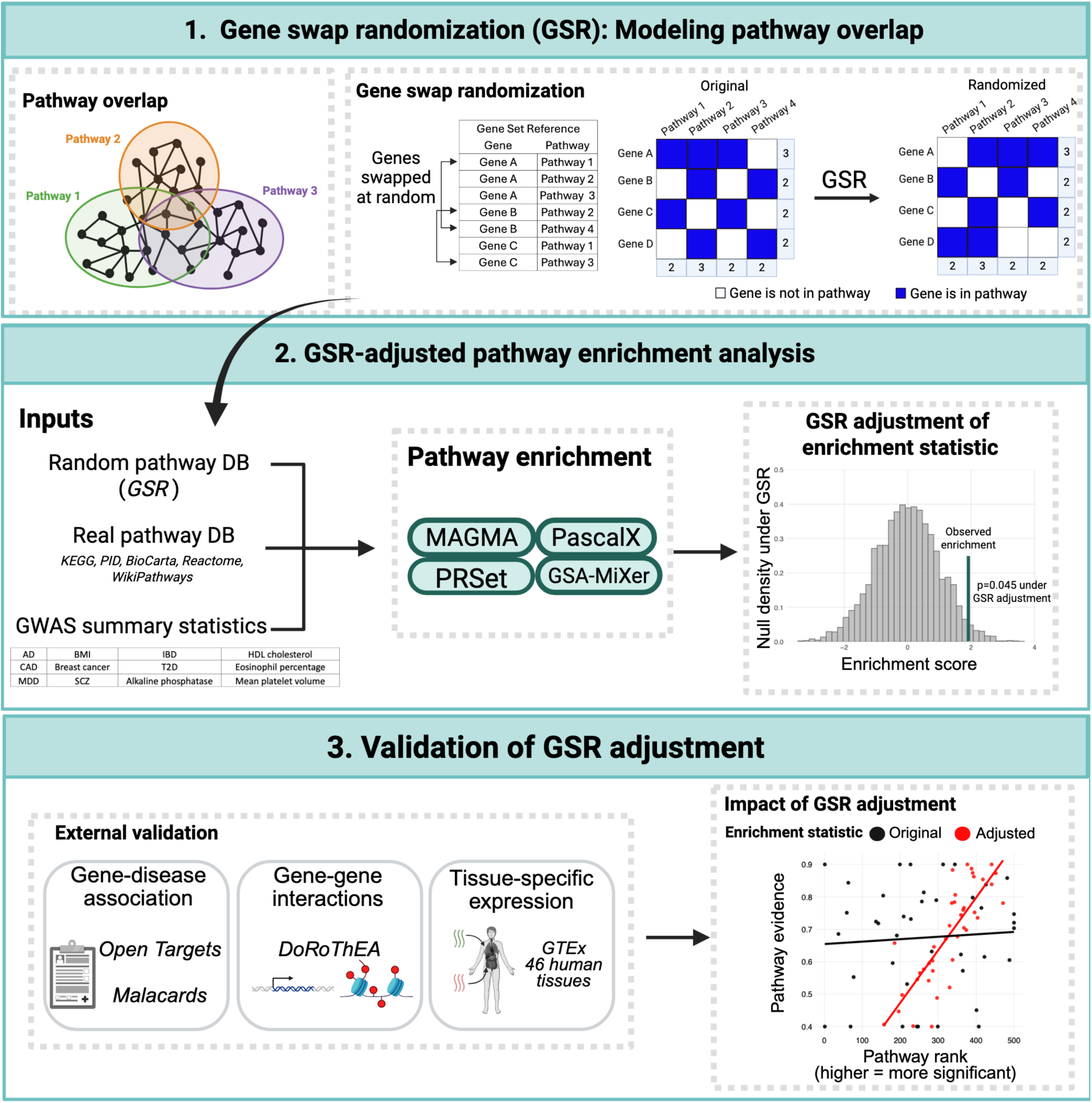
Flowchart of study design.

## Results

### Overview of Gene Swap Randomization (GSR)

Here we introduce Gene Swap Randomization (GSR), which employs a Monte Carlo “switching algorithm” to produce an empirical null set of random pathways of the same size, gene content and pathway overlap as a real set of pathways (e.g. from a canonical database, such as KEGG or Reactome). In each iteration of GSR, two gene entries in the pathway database are chosen at random, and their pathway assignments are swapped, provided the swap does not introduce duplicate genes within a pathway (**Fig 1**). GSR incorporates pathway overlap directly into the null model of a pathway enrichment test, enabling direct adjustment of enrichment statistics. Importantly, GSR retains the observed gene-phenotype associations, which aligns with the desired null hypothesis for GWAS: that gene-level associations are fixed but pathway structure is exchangeable (see Methods for more details). GSR requires only GWAS summary statistics and pre-defined pathways and is therefore broadly applicable.

### Characteristics of real and random pathways

We used MSigDB as our reference pathway database, given its widespread use in GWAS pathway enrichment analyses^15–18^. Across MSigDB, pathway sizes follow a positively skewed distribution, with most pathways containing few genes (median size=37) and a long tail of pathways comprising many hundreds of genes. Gene membership is similarly uneven; most genes appear in one or several pathways and a subset of genes recur across many pathways, which we refer to as “multi-pathway” genes (**Fig S1**). For example, 2,588 genes occur in more than 50 pathways, and 591 genes occur in more than 100 pathways.

As pathway size increases, the number of multi-pathway genes present in the pathway increases, while their proportional representation decreases (**Fig S2**). This pattern is observed both in real MSigDB pathways and random pathways generated via GSR, although the negative correlation between pathway size and the proportion of multi-pathway genes is substantially weaker under randomization. Consistent with this structure, we observe a strong negative correlation between a gene’s frequency in the pathway database and the average size of the random pathways to which it is assigned under GSR: genes that appear in many pathways are, on average, assigned more often to smaller random pathways (**Fig S3**). This behavior is expected given the highly skewed pathway size distribution, characterized by many small pathways and fewer large ones, and the constraints imposed by GSR, which simultaneously preserves pathway sizes and gene-level pathway membership frequencies. We highlight these patterns as relevant context for interpreting the enrichment results below.

### Modeling pathway overlap reveals gene frequency-dependent inflation in enrichment results

To isolate the effect of pathway overlap on pathway enrichment results, we evaluated the behavior of pathway enrichment statistics under controlled null conditions using randomized pathways. Specifically, we performed GWAS pathway enrichment using randomized pathways constructed under two null models: 1) gene set randomization (GSR), which preserves the gene overlap structure across pathways, and 2) randomization preserving pathway size only (PS), which is more aligned with standard gene randomization approaches to enrichment testing. For each pathway, 1,000 randomized pathways were generated under each null model. We estimate nominal type 1 error rates under the GSR and PS nulls and interpret any excess of significant pathway associations as primarily reflecting structural properties of the pathway database rather than genuine pathway enrichment.

We applied several widely used pathway enrichment approaches that differ in their statistical assumptions and data requirements. MAGMA^9^, a popular method for GWAS pathway analysis, performs competitive gene set testing using a two-step, flexible regression framework. Pascal^19^, like MAGMA, using a two-step approach, estimating LD-corrected gene scores and then combining independent and correlated (“fusion”) genes within pathways using a non-permutation-based test. PRSet^20^ computes pathway-specific polygenic risk scores using individual-level data and tests their association with phenotypes. Although its authors do not formally recommend PRSet as an enrichment tool over established methods due to low power in small target samples, PRSet generates pathway-level significance estimates and is therefore included here for comparison alongside more established enrichment approaches. Lastly, GSA-MiXeR^21^ is a recent extension of the MiXeR framework that estimates fold enrichment of partitioned heritability attributable to a gene set. This tool does not produce formal p-values, preventing direct false positive rate estimation.

Several GWAS traits exhibited an excess of nominally significant pathway enrichment when null pathways were generated using GSR (**Fig 2**). Overall, we observed ten GWAS trait-enrichment method combinations with a mean false positive rate (FPR) exceeding 0.10 under GSR, compared to only five such combinations under PS randomization. Under PS randomization, inflation was observed only for PRSet, whereas Pascal appeared well calibrated, and MAGMA even showed slight deflation relative to expectation. In contrast, under GSR, PRSet exhibited the greatest inflation (FPR range: 0.0452-0.414), followed by Pascal (0.0474-0.144), then MAGMA (FPR range 0.0279-0.0836). To assess the variability of FPR across pathways, we examined the 95% middle distribution (2.5th-97.5th percentile) of pathway-level FPR values for each trait. Under PS randomization, this range was narrow and centered near the nominal 0.05 threshold for MAGMA and Pascal, with slight deflation for MAGMA, indicating relatively consistent FPR control across pathways for both tools. In contrast, GSR produced substantially wider and more elevated ranges, with the 95% distribution often exceeding 0.05, particularly for Pascal and PRSet.

**Figure 2.**
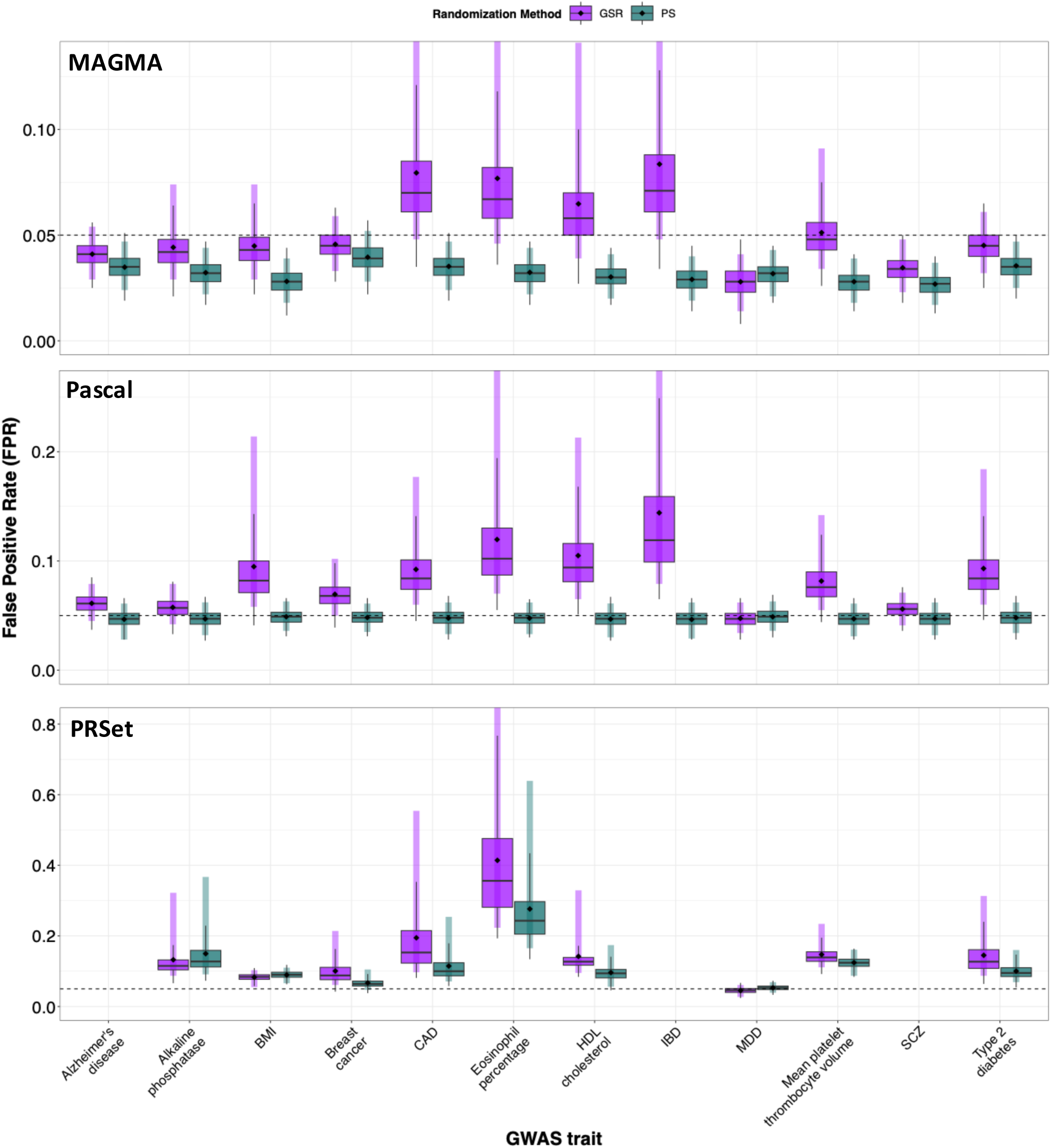
Pathway enrichment statistics show inflation under gene frequency-preserving nulls. False positive rate (FPR) for pathway enrichment analysis as estimated from null models using two randomization approaches: gene set randomization (GSR) and pathway-size preserving randomization (PS). FPR estimated per pathway across 12 GWAS traits using three enrichment methods: MAGMA, PRSet, and Pascal. Boxplots show the distribution of FPR values estimated across pathways for each trait. PRSet excludes Alzheimer’s (AD), inflammatory bowel disease (IBD), and schizophrenia (SCZ) due to limited target sample size (see Methods). Diamonds indicate mean values. The horizontal dashed line indicates the α = 0.05 significance threshold. Colored vertical lines represent the 2.5^th^ to 97.5^th^ percentile range (middle 95% of the FPR distribution). For visualization purposes, some percentile ranges extend beyond the plotted y-axis limits but are not shown.

In summary, under the GSR null we observe both elevated mean FPR and substantial heterogeneity in pathway-level FPR for some GWAS traits and enrichment methods, with particularly pronounced effects for coronary artery disease (CAD), eosinophil percentage, HDL cholesterol, and inflammatory bowel disease (IBD). These results demonstrate that structural features of the pathway database (multi-pathway genes and variation in pathway size) influence the null distribution of enrichment statistics. Enrichment statistics are therefore sensitive to the modeling of pathway overlap in the null, and this feature is not explicitly incorporated into existing enrichment methods. This suggests that, in real pathway analyses, pathway-level significance may be influenced not only by biological signal but also by structural properties of pathway annotations.

For each trait, we also generated QQ plots comparing observed pathway enrichment p-values to the empirical null distributions derived from the 1,000 permutations under each null model (GSR and PS) (**Fig 3; Fig S4-6**). For each null, we summarize the empirical distribution by plotting the median −log10(p-value) at each rank position, allowing a global comparison of observed enrichment patterns against null expectations across the full ranking. Across many trait-method combinations, the highest-ranked observed pathways exceed null expectations under both null models, indicating that top pathways show evidence of enrichment beyond chance. We also observe that the GSR null yields elevated expected −log10(p-values) relative to the PS null. This indicates that the GSR null provides a stricter assessment of pathway significance than pathway-size-only randomization.

**Figure 3.**
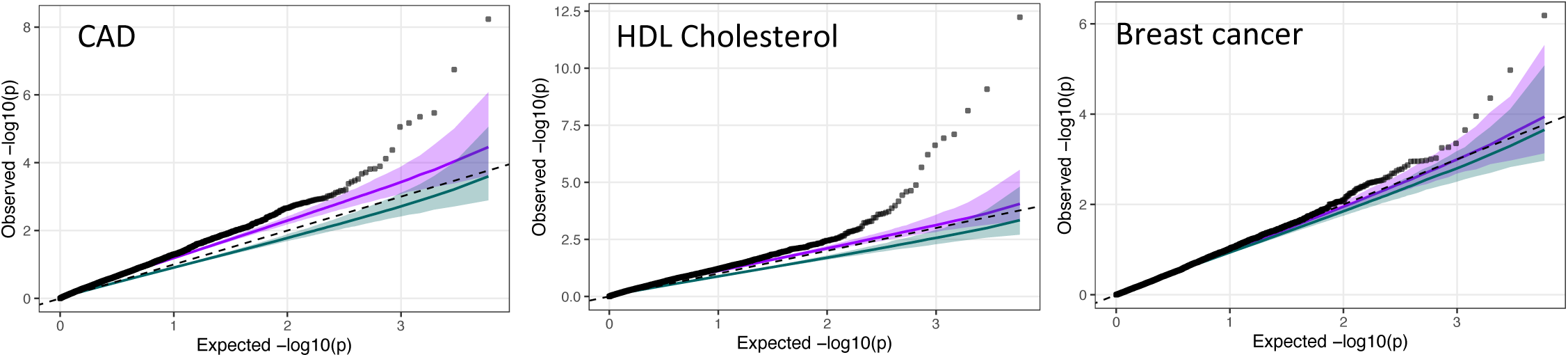
Example QQ plots comparing observed MAGMA pathway enrichment p-values (black points) to two null models: GSR (purple) and PS (green), across multiple traits. Lines show median –log10(p-value) at a given rank position, and shaded regions show 2.5%-97.5% percentile bands at a given rank position (95% of the 1000 permuted p-values at the same rank fall within the shaded region). The dashed line indicates the expectation under a uniform null distribution.

Next, we tested whether the observed inflation under GSR was associated with pathway size. Across many traits and enrichment methods, we observed a strong positive relationship between pathway size and the degree of inflation, such that the excess of nominally significant pathways increased with pathway size (**Fig 4; Fig S7-9)**. Large random pathways were therefore more likely to be declared significantly enriched under the GSR null model. This size-dependent inflation was not observed when using PS-based randomization. The effect was not explained by GWAS trait heritability or SNP polygenicity (**Fig S10-11**). Inflation was strongest for traits whose associated genes were enriched for multi-pathway genes (**Fig 5**), consistent with the expectation that preserving gene frequency in the null model has the greatest impact when multi-pathway genes are themselves enriched for trait association. These results have important implications for real pathway analyses. If the GSR null is assumed to be appropriate, then large real pathways will also be more likely to appear significantly enriched. Together, these results suggest that preserving gene membership frequency in the null exposes sensitivity of enrichment tests to pathway database structure, particularly for large pathways, and highlights the importance of distinguishing pathway-specific enrichment from enrichment driven by recurrent gene sharing across pathways.

**Figure 4.**
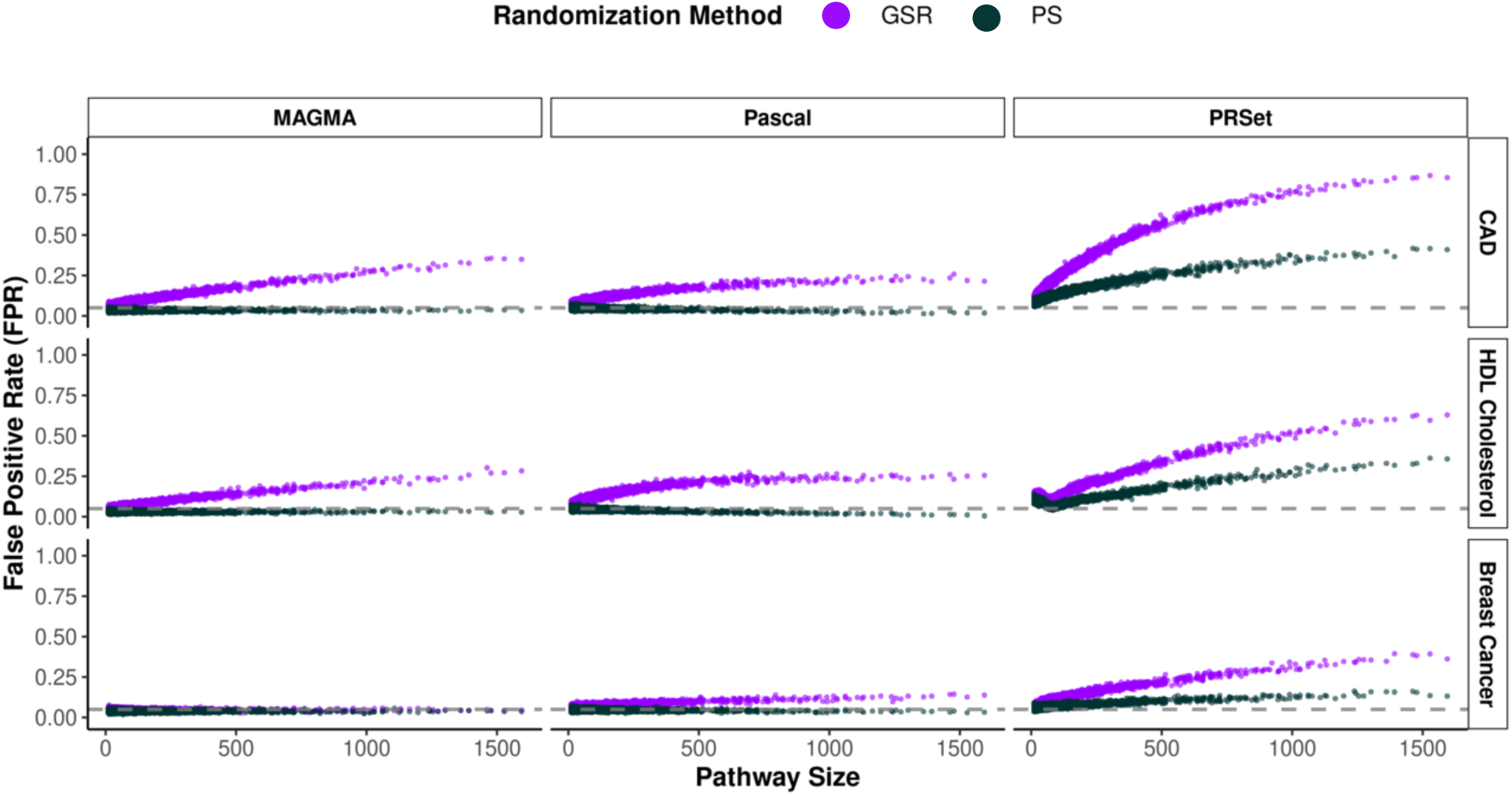
Inflation with GSR null is dependent on pathway size. Relationship between pathway size and estimated FPR for three representative traits: coronary artery disease, HDL cholesterol, and breast cancer.

**Figure 5.**
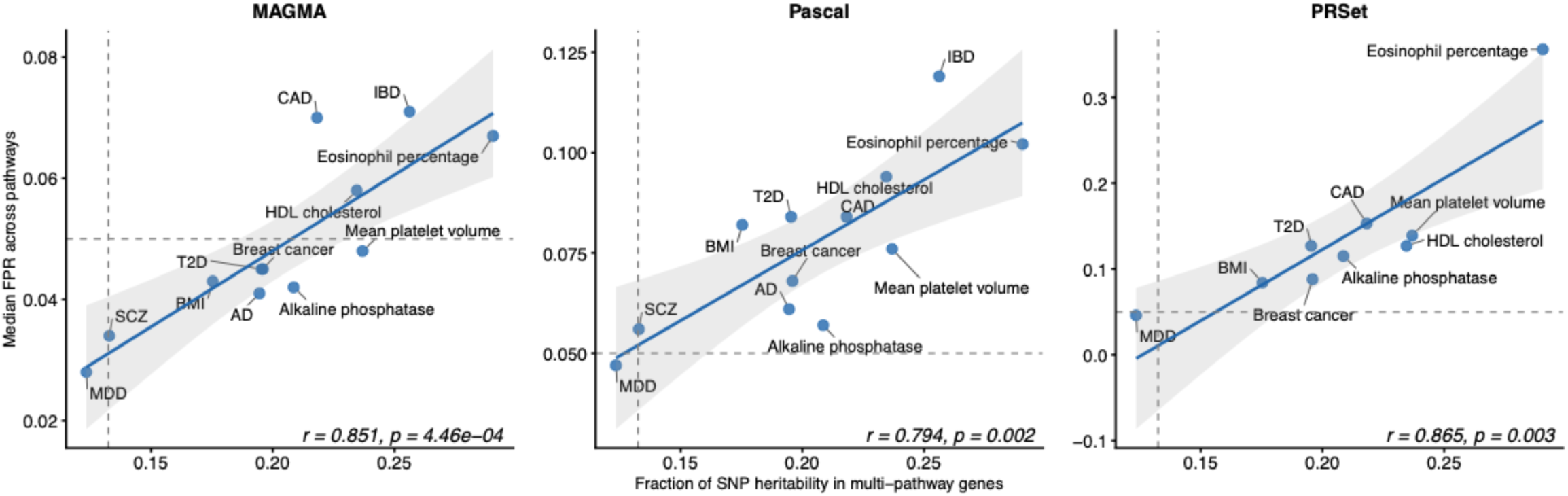
Relationship between multi-pathway gene heritability enrichment and false positive rate using the GSR null model across traits and enrichment tools. Scatter plots show for each trait the median false positive rate (FPR) across pathways against the fraction of SNP heritability attributable to multi-pathway genes using stratified LD score regression and normalized relative to all genes in the pathway database, defined as 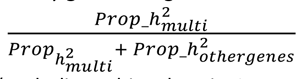.Each panel corresponds to a pathway enrichment tool: MAGMA, Pascal, and PRSet (excluding schizophrenia, IBD, and AD). The vertical dashed line denotes the null expectation under no heritability enrichment, i.e. the fraction of SNPs residing in multi-pathway genes. The horizontal dashed line marks FPR = 0.05.

### Adjustment for pathway overlap improves prioritization of trait-relevant pathways

Because GWAS enrichment analyses often yield multiple significant pathways, the practical benefit of these analyses typically depends on the accurate prioritization of the most biologically important pathways for downstream follow-up. For this reason, we next evaluated whether adjusting GWAS pathway enrichment results with respect to the GSR null model, through the calculation of GSR-derived empirical p-values and standardized effect sizes, improves the accuracy of prioritized pathways. Our hypothesis was that accurate reranking of pathway enrichment results will better prioritize pathways with external evidence of disease association, gene regulatory relationships, and/or expression in a trait-relevant tissue. We compared pathway rankings obtained under standard use of these pathway analysis tools to those obtained after GSR-based adjustment of the test statistics. We compared pathway rankings to four external validation resources: Open Targets gene-disease associations^22^, Malacards gene-disease associations^23^, DoRothEA transcription factor-target gene interactions^24^, and scores of tissue-specificity of gene expression derived from the Genotype Tissue Expression project (GTEx)^25,26^. We estimated the Spearman correlation between pathway rankings and these external validation scores. We examine only the rankings of the top 500 pathways (as defined by the original, unadjusted p-value for each tool), with the understanding that most tested pathways are not significantly associated with a given GWAS trait. Given the absence of p-values in the GSA-MiXeR tool, we instead define GSA-MiXeR top pathways as the union of top 500 pathways across MAGMA, Pascal, and PRSet results for a given GWAS trait.

Given our understanding of how pathway size interacts with inflation under the GSR null, we first examined the relationship between external evidence scores and pathway size. Evidence scores were strongly pathway-size dependent, with large values confined to small gene sets and both variance and magnitude decreasing as pathway size increases (**Fig S12-15)**. This is important to note because the GSR null model tends to upweight smaller pathways and downweight larger ones, particularly for MAGMA, PRSet, and Pascal (**Fig S16-19**). Interestingly, for MAGA, PRSet, and Pascal, original enrichment rankings were associatd with pathway size, with larger pathways more likely to be prioritized (**Fig S20**). Following GSR adjustment, this association was attenuated, indicating that pathway size influences baseline rankings but is mitigated under the GSR null. Taken together, these results suggest that GSR adjustment can shift pathway rankings in a manner that reflects size-dependent structure present in both enrichment statistics and external validation resources. At the same time, smaller pathways are often more functionally coherent, whereas larger pathways frequently represent higher-level aggregations of related biological processes^27^. These considerations underscore that concordance between pathway rankings and external evidence should be interpreted in the context of pathway size.

Across most GWAS traits, enrichment methods, and validation metrics, pathway rankings show positive Spearman correlation with external evidence scores, indicating that these approaches generally prioritize biologically relevant pathways (**Fig 6**). Differences between rankings before and after GSR adjustment were largest when evaluated against Open Targets and Malacards gene-disease resources, whereas tissue specificity tests and DoRothEA showed appreciable changes with GSR primarily for PRSet.

**Figure 6.**
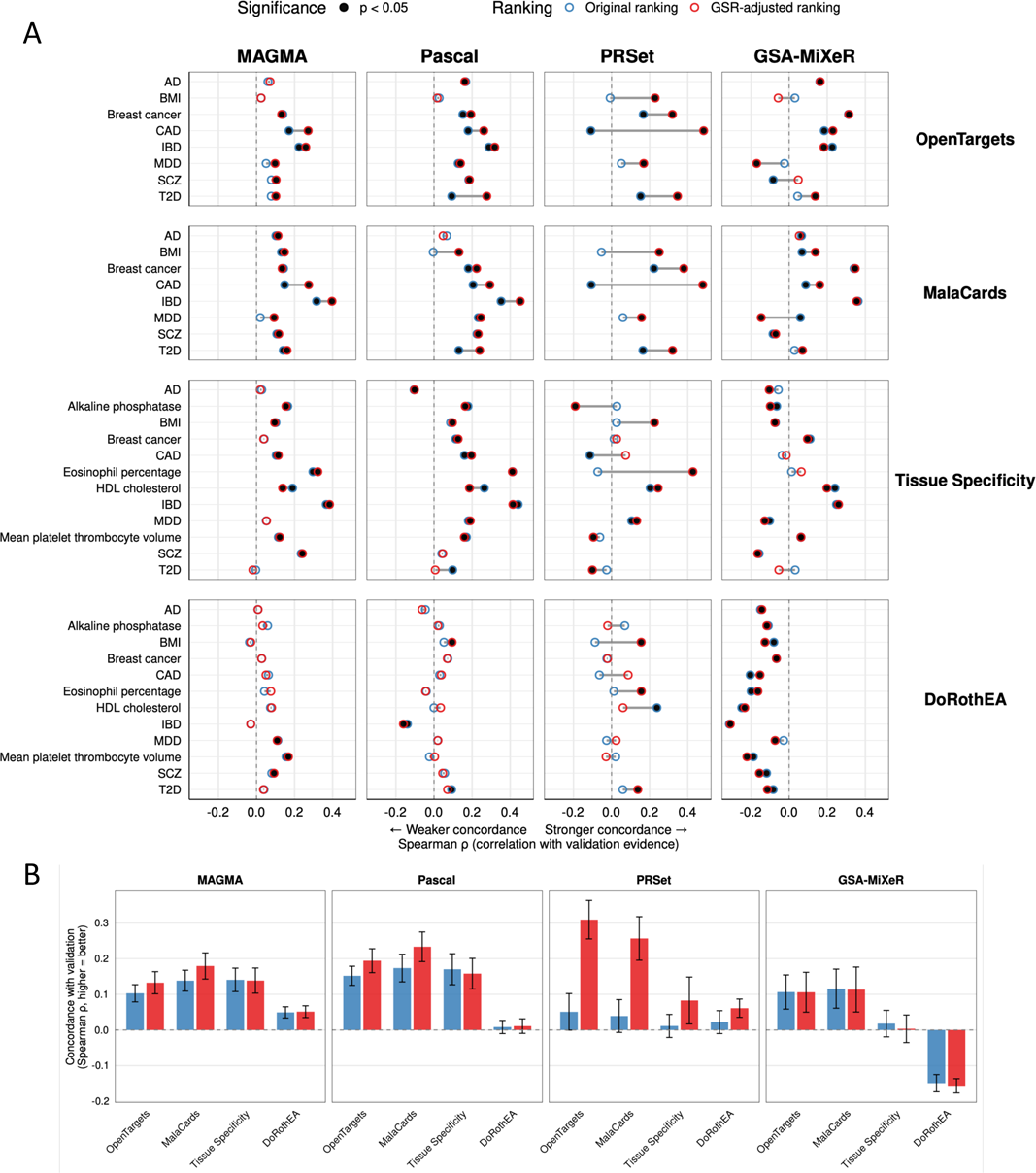
Impact of GSR adjustment on concordance between GWAS pathway rankings and external evidence. A) Spearman correlation between pathway rankings and external validation evidence before (blue outline) and after (red outline) GSR adjustment for top 500 pathways. Positive correlations indicate better alignment with validation evidence. Lines connect before/after values for each trait. Each panel represents a different combination of pathway analysis method (columns: MAGMA, Pascal, PRSet, GSA-MiXeR) and validation framework (rows: OpenTargets disease-gene associations, MalaCards disease-gene associations, tissue-specific gene expression, and DoRothEA transcription factor targets). Points filled black when correlation reaches nominal significance (p<0.05).\ PRSet results for schizophrenia (SCZ), Alzheimer’s disease (AD), and inflammatory bowel disease (IBD) are excluded due to limited target sample size. B) Mean Spearman correlations (ρ) for top 500 pathways across the four validation frameworks. Error bars indicate standard error of the mean correlation across traits. Note that all correlations are considered in this plot regardless of statistical significance.

For MAGMA and Pascal, GSR adjustment improved concordance with external validation metrics across traits and validation sources. These improvements were most evident for CAD, IBD, and T2D, when examined using Open Targets and Malacards references (**Fig 6**). Improvements were small in magnitude but consistent in direction, indicating that adjustment refines pathway prioritization rather than substantially reordering rankings. Importantly, negative correlations, where adjustment reduced concordance, were less common and of smaller magnitude.

In contrast, PRSet showed larger positive shifts in performance following GSR adjustment with greatest improvements observed for eosinophil percentage, CAD, T2D, breast cancer, and BMI. Results for GSA-MiXeR were more heterogeneous: GSR adjustment improved concordance with external evidence for some traits, but in other cases rankings showed negative correlation, regardless of whether GSR adjustment was used, particularly when evaluated against DoRothEA or tissue-specific expression. In the original GSA-MiXeR study, the authors found the method performed well when ranking pathways pre-filtered to those deemed significant by MAGMA. Perhaps the incorporation of other tools, especially those exhibiting inflation such as PRSet, introduced non-significant pathways that degrade GSA-MiXeR ranking performance. Developing strategies for pathway ranking in enrichment methods that do not provide formal significance testing like GSA-MiXeR remains an important direction for future work.

In summary, ranking enrichment results by empirical p-values and standardized effect sizes derived from our null model, accounting for both gene frequency and pathway size, yielded overall stronger concordance with external evidence scores, with most impact on results of PRSet, Pascal, and MAGMA methods.

## Discussion

Pathway enrichment analysis is routinely applied in post-GWAS studies to translate variant-level associations into biological mechanisms, and the interpretation of enrichment statistics therefore has broad implications for genetic association results. However, pathway databases contain extensive gene sharing across pathways that is not explicitly modeled by standard enrichment nulls. Here, we show that pathway overlap alone can generate enrichment under polygenicity, even when there is no pathway-specific signal, particularly for large pathways and traits enriched for multi-pathway genes. We introduce Gene Swap Randomization (GSR), a pathway overlap-aware null model that conditions on gene frequency and pathway size, allowing for the explicit modeling of multi-pathway genes while preserving gene-trait associations. Across 12 diverse GWAS traits and four pathway tools (MAGMA, Pascal, GSA-MiXeR, PRSet), we show that accounting for pathway overlap refines pathway ranking and improves concordance with external disease and functional benchmarks. These results indicate that the structural properties of pathway databases meaningfully influence GWAS enrichment results and should be considered in their interpretation.

Our study further emphasizes that GWAS pathway enrichment statistics depend critically on the choice of null model. When we test the enrichment of random pathways that preserve only pathway size, statistics appear well-calibrated for most tools. However, under the GSR null, which additionally preserves gene frequency across pathways, enrichment of random gene sets exceeds the nominal 5% threshold in many traits. This inflation increases with pathway size and is most pronounced for traits in which multi-pathway genes are themselves enriched for trait association. These results demonstrate that pathway overlap affects null expectations and that commonly used enrichment tools are sensitive to the modeling of multi-pathway genes in the null. Consequently, pathway significance may partly reflect structural properties of the pathway database rather than pathway-specific biology, particularly when GWAS signal is concentrated in genes that appear across many pathways. We therefore recommend that researchers evaluate the distribution of gene association as a function of gene frequency and apply GSR adjustment when multi-pathway genes show strong association with the GWAS trait (**Fig 5**).

Related to this, we observed that for MAGMA, PRSet, and Pascal, pathway enrichment rankings were associated with pathway size, with larger pathways more likely to be identified as significant. This relationship was attenuated following GSR adjustment, indicating that pathway size contributes to baseline enrichment rankings but that this effect is partially mitigated when multipathway genes are explicitly modeled. The impact of pathway size on enrichment results is multifactorial and not solely driven by pathway overlap^28,29^, and the true relationship between biological signal and pathway size remains unclear. Still, the attenuation of size-dependent ranking patterns following GSR adjustment is encouraging and suggests that GSR can help reduce systematic biases and improve pathway prioritization.

Consistent with these shifts in ranking behavior, GSR adjustment improved the biological prioritization of pathways across most enrichment methods. Reranking pathways using GSR-derived empirical p-values and standardized effect sizes increased concordance with external validation benchmarks for MAGMA, Pascal, and PRSet in a majority of trait comparisons. Improvements were most pronounced for disease-associated benchmarks, whereas functional genomics benchmarks showed weaker sensitivity across traits, with many exhibiting little or no correlation with validation scores irrespective of adjustment. PRSet showed the largest improvements following GSR adjustment. Because PRSet aggregates individual-level polygenic signal and is optimized for prediction, results may be disproportionately affected by variants with stronger effects on the GWAS trait. As a result, multi-pathway genes that harbor strong GWAS signals can drive enrichment across multiple pathways. Although this property is advantageous when PRSet is used for trait or subtype prediction, it complicates interpretation when pathway results are used to infer disease mechanisms. Results were more variable for GSA-MiXeR, which models fold enrichment rather than pathway-level significance and does not produce formal p-values. In contrast to the original GSA-MiXeR evaluation, where prioritization was restricted to MAGMA-significant pathways, we considered the broader union of top pathways across methods, potentially introducing pathways with limited enrichment under this model.

We highlight several limitations of this study. First, GSR is intended as a pathway prioritization tool rather than a formal hypothesis-testing procedure with guaranteed error control. Because empirical p-values are calculated separately for each pathway under a shared randomization scheme that preserves pathway size and gene frequency, GSR does not provide formal family-wise error rate control across pathways. Instead, it quantifies the extent to which a given pathway’s enrichment exceeds what would be expected given the structure of the pathway database. GSR-derived p-values are therefore suitable for pathway prioritization but not to replace standard p-values for inference. Second, like many gene randomization approaches, GSR null pathways are not matched to real pathways in terms of gene-gene correlation. This can lead to false positives if genes with dense LD patterns drive associations^1^. However, the enrichment methods evaluated here each incorporate internal models of LD, which likely partly mitigate this concern. Future extensions of GSR could improve adjustment by modeling gene-gene correlation or LD-aware matching. Finally, we focused on the MSigDB reference due to its popularity in GWAS enrichment studies; patterns of gene frequency and pathway overlap may differ in other databases and warrant further evaluation.

This is to our knowledge the first study to systematically evaluate how gene frequency and pathway overlap influence GWAS pathway enrichment. By constructing an empirical null that preserves both pathway size and gene frequency, we show that multi-pathway genes can induce pathway size-dependent inflation and affect pathway prioritization in widely used enrichment tools. GSR models this structure and improves biological prioritization of top pathways without replacing existing enrichment tools, providing a generalizable framework for pathway overlap-aware adjustment of enrichment statistics. As pathway databases continue to expand in size and redundancy, accounting for multi-pathway genes will be increasingly important for robust interpretation of enrichment results, and we suggest that pathway overlap-aware null models be considered in future methodological development.

## Methods

### Gene pathway reference

We obtained Reactome, KEGG, BioCarta, WikiPathways, and Pathway Interaction Database (PID) pathways from the Molecular Signatures Database (C2) curated gene sets (MsigDB v2023)^30^. We converted Gene IDs to Ensembl IDs, and filtered for autosomal, protein coding genes. We removed pathways with fewer than 10 genes or more than 2,000 genes. The final pathway database file included 16,605 genes across 5,878 pathways.

### Genomic swap randomization

To simulate pathway enrichment under a null model that preserves both pathway size and the frequency of genes across pathways, we employed a Monte Carlo “switching algorithm”, also known as swap randomization. In each iteration, two gene entries in the pathway database are chosen at random, and their pathway assignments are swapped, provided the swap does not introduce duplicate genes within a pathway (**Fig 1**). This process can also be visualized as a binary event matrix, where rows are the total number of unique genes, and columns are the pathways. Through swap randomization, the row sums (gene frequency) and column sums (pathway size) are fixed, and gene-pathway connections are shuffled at random. Similarly, we can visualize a pathway database as a bipartite network, with one set of nodes representing genes and the other representing pathways, and all network connections occur only between these two sets of nodes.

We determined the number of required swaps using the empirical lower bound proposed by Gobbi et al. (2014)^31^. Considering the pathway database as a bipartite network, Gobbi et al. proposes an approximate minimum number of required swaps equal to

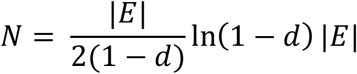

where |*E*|is the number of existing links (i.e. gene-pathway relationships) and *d* is the ratio between |*E*| and the number of links of a fully connected bipartite network with the same number of nodes in each set: 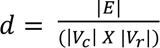. Here *V_r_* represents the number of unique genes, and *V_c_* represents the number of pathways. By implementing the switching algorithm with this empirical lower bound, each randomized version of the pathway database reaches two conditions:

1. the average similarity between the original and randomized database remains constant as the number of switching steps increases, and
2. the average similarity of the original database and the randomized database is comparable to the similarity between any two randomized databases with the same number of genes, pathways, and gene-pathway connections.

We implemented the algorithm using the *BiRewire* v3.30 R package to generate 1,000 randomized pathway databases^31^. Default parameters were used (accuracy = 0.00005, max.iter = “n”), where the number of switching steps to be performed corresponded to the lower bound defined in Gobbi et al.

### Trait selection

This study included published GWAS for eight traits: body mass index (BMI)^32^, coronary artery disease (CAD)^33^, type 2 diabetes (T2D)^34^, major depressive disorder (MDD)^35^, Alzheimer’s disease (AD)^15^, schizophrenia (SCZ)^36^, inflammatory bowel disease (IBD)^16^, and breast cancer^37^. We selected the largest available GWAS of genotype-derived European ancestry that do not include UK Biobank data (UKB), to ensure we are able to test *PRSet* performance with UKB target data without facing issues of sample overlap. In addition, we conducted GWAS of four lab measurements in the UK Biobank: HDL cholesterol, mean platelet thrombocyte volume, alkaline phosphatase, and eosinophil percentage. These 12 total traits were selected to provide a balance of continuous and dichotomous traits across diverse domains, including anthropometric, cardiometabolic, central nervous system, immune, and cancer-related phenotypes (**Table S1**).

#### UK Biobank GWAS

The UK Biobank is a prospective cohort of about 500,000 individuals, recruited at ages 40-69 years, to study long-term health and disease. Participant data include genetic, biomarker, healthcare, physical, imaging, and questionnaire data collected at baseline and/or follow-up.

The genetic dataset consists of 488,377 samples genotyped at 805,426 SNPs. Using 4-means clustering analysis on the first two principal components of the genotype data and projection onto 1000 Genomes reference populations, we identified 461,788 individuals of European ancestry. Standard genotype QC was then applied, removing SNPs with a minor allele frequency < 0.01, genotype missingness > 0.02, and Hardy-Weinberg equilibrium test p-value < 10^-8^, and restriction to autosomes. We further excluded 72,808 related individuals (kinship coefficient > 0.044), and individuals who had a high degree of missingness or heterozygosity, or mismatched genotype-derived and self-reported sex, leaving 387,235 individuals and 542,312 SNPs.

We next selected blood measurement traits of interest, considering 60 lab values in total. SNP-heritability estimates for these traits were obtained from the Neale Lab (https://nealelab.github.io/UKBB_ldsc/), calculated in the UK Biobank using partitioned LD score regression with the MTAG implementation of LDSC^38–40^. We filtered for traits with SNP-heritability estimates (h^2^) greater than 0.1 and p-values less than 0.05. Next, we calculated Pearson correlations among remaining lab values and performed clumping. Starting with the trait with the highest heritability estimate, we iteratively removed all correlated traits (|r| > 0.1), yielding a set of ten independent lab values. We finally selected four representative lab values: HDL cholesterol, mean platelet thrombocyte volume, alkaline phosphatase, and eosinophil percentage, to test across traits with diverse biological underpinnings and reasonable heritability.

GWAS were performed in PLINK 2.0 using the --glm function for linear regression, adjusting for age, sex, center, and the first 10 genotype-derived ancestry PCs^41^. We randomly split the sample, using 70% for discovery GWAS and holding out 30% as a target dataset for later *PRSet* analyses (**Table S2**).

### Stratified LD score regression analysis

#### Gene set definition and annotation

Pathway membership was defined as the number of gene sets in which a gene appeared. Genes at or above the 90^th^ percentile of this distribution were classified as multi-pathway genes. SNPs were assigned to genes within a symmetric ± 35 kb window. Binary SNP annotations for multi-pathway and non-multi-pathway gene groups were generated using make_annot.py from the LD Score Regression (LDSC) software, based on the 1000 Genomes Project Phase 3 European reference panel.

LD scores were computed with a 1cM window, retaining HapMap3 SNPs. Baseline LD annotations (v2.2), along with precomputed allele frequencies and regression weights, were included. GWAS summary statistics were harmonized using munge_sumstats.py, with SNPs restricted to HapMap3 variants and aligned to the reference panel.

#### Partitioned heritability analysis

Stratified LD score regression (s-LDSC v1.0.1) was applied to estimate the contribution of multi-pathway genes to SNP heritability. Models included baseline LD annotations (v2.2), multi-pathway genes, and other pathway database genes, with –overlap-annot enabled. We quantified the contribution of multi-pathway genes to SNP heritability as:

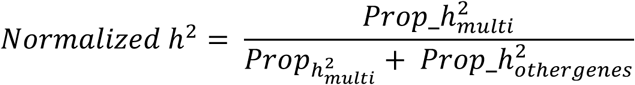

which represents the fraction of SNP heritability attributable to multi-pathway genes, relative to the total heritability captured by genes in the pathway database. Because *Prop*_ℎ^2^ values are defined relative to total SNP heritability, this normalization is equivalent to 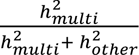

GWAS gene set enrichment

We performed competitive GWAS pathway enrichment tests across these 12 traits using four methods: *MAGMA, PRSet, GSA-MiXeR,* and *PascalX*. While not explicitly developed as a gene set enrichment tool, *PRSet* is included due to the increasing popularity of pathway-based PRS to identify relevant pathway biology underlying GWAS associations. The 1,000 randomized pathway databases, as well as the real pathway database, were provided as input to each method.

#### MAGMA (Multi-marker Analysis of GenoMic Annotation)

We applied *MAGMA* (v1.10) using the SNP-wise mean base model to perform competitive gene set enrichment analysis using GWAS summary statistics. The *MAGMA* tool utilizes a flexible multiple regression and a two-step approach to gene set enrichment. First, variant-level associations are aggregated at the gene level, using the sum of squared SNP Z-statistics as the test statistic, reflecting the strength of each gene’s association with the phenotype. SNPs were mapped to genes using a symmetric ± 35 kb window. Second, these gene-level associations are modeled as the dependent variable in a multiple regression framework calculated as:

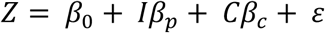

where *I* is a binary variable that takes the value of 1 or 0 indicating if a gene is in the pathway *p*, and *C* is a matrix of covariates (default covariates include log transformed values of gene size, gene density, sample size, and inverse minor allele count). The competitive gene set analysis tests the null hypothesis *β_p_* = 0 against the one-sided alternative hypothesis *β_p_* > 0, determing whether genes within the gene set show stronger association than genes outside the set.

#### PRSet: Pathway-based polygenic risk scores

*PRSet* is a software to calculated polygenic risk scores (PRS) across biological pathways per individual and test their association with a trait^20^. *PRSet* applies clumping and thresholding^42^ on each pathway to generate *k* PRSs relating to *k* pathways for each individual *i*:

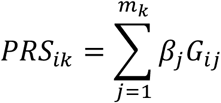

where *m_k_* is the number of clumped SNPs in pathway *k*, *β*_j_ is the GWAS weight for SNP *j*, and *G_i_*_j_ is the genotype of individual *i* for SNP *j*. To assess significance of pathway association while controlling for pathway size, *PRSet* performs permutation testing, generating random pathways with the same number of SNPs as the test set and calculating an empirical “competitive” p-value:

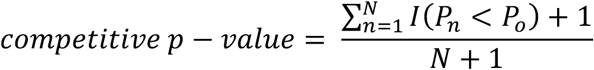

where *I* takes a value of 1 if the observed p-value *P_o_* is larger than the one obtained from the *n*th null set *P_n_*.

We calculated pathway PRSs, assigning SNPs to genes within a 35 kilobase window, and including all SNPs mapped to each pathway (GWAS p-value threshold = 1). A UK Biobank subsample was used as individual-level target data, to test association between pathway PRSs and the trait. Prior simulation work shows that PRSet has limited performance in certain settings when applied to small target samples (n=1,000)^20^. We therefore restricted our analyses to traits with more than 2,000 individuals available in the UK Biobank target sample (or more than 2,000 cases for dichotomous traits). This excluded Alzheimer’s disease, inflammatory bowel disease, and schizophrenia from PRSet analyses. To optimize the runtime, we first adjusted each UKB outcome for patient sex, age, sequencing batch, center, and the first 15 genotype-derived PCs and obtained the residuals. Then we tested for association between outcome residuals and pathway PRSs. 1,000 permutations were performed to determine competitive p-values.

#### GSA-MiXeR

*GSA-MiXeR*, an extension of the *MiXeR* framework^43^, is a competitive pathway enrichment method that measures the fold enrichment of partitioned heritability attributable to a gene set^21^. *MiXeR* models SNP effects using a spike-and-slab prior, where most variants have no effect and a subset of variants have normally distributed nonzero effects. In *GSA-MiXeR* two models are fit to the GWAS summary statistics: a full model, where each gene has its own estimated contribution to heritability, and a baseline model where all genes are assumed to contribute equally. Fold enrichment is calculated as the ratio of a gene set’s heritability under the full vs the baseline model, after accounting for LD, allele frequency, and functional annotations:

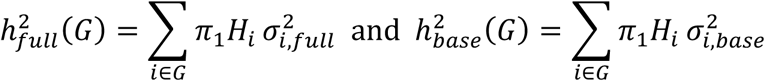

where *π*_1_ is the prior probability of a variant having an effect, *H_i_* is the heterozygosity of the *i*th SNP, and *σ*^2^ is the effect size variance of the *i*th SNP. Unlike some other enrichment tools, GSA-MiXeR estimates an effect size (fold enrichment) rather than a p-value, allowing for the comparison of enrichment results across pathways. Further details can be found at Frei et al (2024).

#### PascalX

*PascalX,* like *MAGMA*, uses a two-step approach to competitive pathway enrichment, estimating first gene-level and then pathway-level association with the GWAS trait^19^. First, SNPs are aggregated to gene-level associations based on proximity (we applied a symmetric 35 kilobase window). SNP Z scores are then used to compute an LD correlation matrix from 1000 Genomes European reference. Gene-level test statistics are calculated using either the maximum-of-chi-squares (MOCS)^44^ or sum-of-chi-squares (SOCS)^45^ statistic, with correction for LD. *PascalX* then applies numeric solutions to derive p-values of MOCS and SOCS statistics, rather than permutations, to improve efficiency.

To account for nearby correlated genes in a pathway, PascalX uses a “fusion gene” approach: all SNPs in a correlated gene cluster (within 1MB by default) are treated as a single gene score, and pathway scores derived from the set of independent genes and fusion genes.

Next, *PascalX* implements a modified Fisher test to calculate pathway statistics, performed in three steps: 1) gene scores are transformed to follow a target distribution, 2) a test statistic is calculated by summing scores across pathway and fusion genes, and 3) empirical methods determine the significance of pathway enrichment. Two options are available for pathway scoring: 1) an empirical sampling method or 2) a chi-squared approach, where all gene p-values genome-wide are ranked 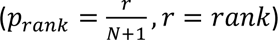, converted to chi-squared scores, and summed across pathway genes.

In our study we applied the SOCS gene statistic and chi-squared pathway statistic approach, as it performed well across datasets in the original validation study^46^.

### Deriving standardized effects and empirical p-values using swap randomization

For a given pathway *X*, we calculate the empirical p-value for enrichment as

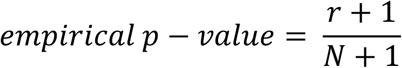

where *N* is the number of randomized versions of pathway *X*, and *r* is the number of those randomized versions with p-values less than the observed p-value for real pathway *X*. Note for *PRSet,* we calculated empirical p-values using our randomized pathways and the unadjusted raw p-values (not the competitive p-values) provided by the software. We add 1 to the numerator and denominator to avoid an empirical p-value of zero and to treat the observed pathway as an additional draw under the null hypothesis^47^. This procedure restricts the smallest empirical p-value to 1/(*N* + 1), which can limit the ranking of highly significant real gene sets.

To allow for more nuanced ranking, we also calculate a standardized effect size (SES)^48^ as

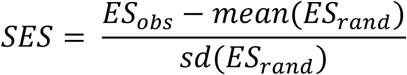

where *ES_obs_* is the observed effect size of pathway *X* and *ES_rand_* represents the effect size of the randomized versions of pathway *X*. Inclusion of a standardized effect size also allows for testing of the *GSA-MiXeR* tool, which does not provide a p-value in the output, and only a measure of effect size (heritability fold enrichment). The SES provides a measure of the observed enrichment effect as compared to our null distribution, accounting for the impacts of gene frequency and pathway size.

### Type 1 error rate estimation

For a given pathway *X*, under each null model the false positive rate (FPR) was defined as the proportion of randomized versions of that pathway with p-values less than 0.05. When viewed in terms of the binary event matrix, this is equivalent to testing the enrichment of a given column (pathway) across *N* randomizations of the matrix and computing the proportion of times it appears significant by chance.

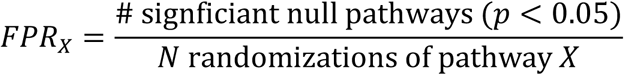

### Benchmarking swap randomization-based reranking of enrichment results

To determine whether reranking pathway enrichment results using our null model improves prioritization of biologically relevant pathways, we compared top-ranked pathways to external datasets of gene-disease relationships (Open Targets, Malacards), gene-gene relationships (DoRothEA), and tissue-specific expression (GTEx). The rationale is that the most effective ranking will prioritize pathways with stronger disease relevance and higher biological confidence (supported by functional evidence). For each GWAS trait and enrichment method, we generated three pathway rankings:

1. By p-value, then effect size (Original ranking)
2. By empirical p-value and standardized effect size using a null model that accounts only for pathway size (PS ranking)
3. By empirical p-value and standardized effect size using the gene swap randomization null model (GSR ranking)

Since most pathways will show non-significant enrichment, and differences between their rankings are not meaningful, we restrict our analysis to only the top 500 pathways for each method. We then calculated Spearman’s correlations between pathway rankings and evidence scores from external datasets. Stronger correlation suggests that the method more effectively prioritizes biologically meaningful enrichment results.

#### Open Targets Comparison

We first compared pathway ranks to the Open Targets Platform dataset of target-disease associations^22^. Direct and indirect associations, by data type, were downloaded from the Open Targets web platform in November 2024 (**Table S2**). Gene-disease scores were filtered to only those based on literature, drug, and animal model evidence, excluding evidence from genetic associations to avoid circularity. Genes without evidence were assigned a score of zero, and then the mean score was calculated for each MSigDB pathway. Open Targets scores were available for eight GWAS traits in the present study: IBD, breast cancer, Alzheimer’s disease, schizophrenia, MDD, T2D, CAD, and BMI (**Table S3**).

#### Malacards Comparison

Gene-disease scores were downloaded from the Malacards Human Disease Database in September 2025 (**Table S3**)^23^. Gene identifiers were converted from HGNC symbol to Ensembl ID. Due to the large range of Malacards scores in the database, scores were rank normalized such that each gene received a score of (*r* + 1)/(*n* + 1), where *r* is the inverse rank of the Malacards gene score and *n* is the total number of genes with scores for that disease. Genes without evidence were assigned a score of zero. For each pathway, we then calculated the mean Malacards score across pathway genes. Malacards scores were available for eight GWAS traits in the present study: Alzheimer’s disease, BMI, breast cancer, CAD (related traits), IBD, MDD, schizophrenia, and T2D (**Table S4**).

#### DoRothEA Comparison

Next we compared pathway ranks to gene-gene relationship evidence from DoRothEA, a curated collection of transcription factor (TF)-target gene relationships from literature review, TF binding motifs, gene co-expression, and ChIP-seq data^24^. We downloaded the full database using the *dorothea* R package v1.10, converted gene identifiers from HGNC symbol to Ensembl ID, and filtered to exclude TF-target gene pairs inferred from literature review. This filtering provided a functionally informed benchmark less affected by literature bias.

For each pathway, we calculated the proportion of genes pairs supported by DoRothEA evidence, either direct TF-target pairs or pairs of target genes regulated by the same TF. Pathways with higher DoRothEA scores contain genes with greater support of shared biological function and can be considered “higher confidence” pathways. We tested whether reranking pathways using our null model increased the prioritization of these higher-confidence pathways.

#### Tissue Specificity Comparison

We next measured association between gene set enrichment rankings and relative (tissue specific) gene expression in tissues with clear biological relevance to each GWAS trait (**Table S5**). For each trait, we chose a single representative GTEx tissue to summarize trait-relevant expression patterns. We note these tissues were used as practical proxies and are not intended to represent the uniquely causal tissue for each trait. For BMI, prior genetic studies demonstrate stronger enrichment of BMI-associated loci in brain tissues relative to adipose tissue, therefore we selected frontal cortex as the representative tissue in this case^49,50^.

Relative expression values were taken from García-González et al. (2026)^25^. Briefly, gene expression from 50 tissues from the GTEx project were downloaded from https://gtexportal.org/home/datasets. First, standard preprocessing steps were applied, including 1) removal of non-protein coding genes or genes with no expression, 2) removing tissues with fewer than 100 samples (45 tissues remained), and 3) scaling the expression such that the total is 10^6^ TPM. Next, tissue-specific expression for each gene *g* in tissue *t* was calculated as the absolute gene expression divided by its total expression across tissues:

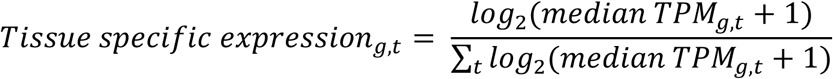

We took the mean tissue specific expression across genes in a given pathway, and tested concordance of these pathway-level scores with gene set enrichment rankings. We tested if reranking results according to our null model resulted in greater prioritization of pathways with trait-relevant tissue-specific gene expression.

## Acknowledgements

This work was supported in part through the computational and data resources and staff expertise provided by Scientific Computing and Data at the Icahn School of Medicine at Mount Sinai and supported by the Clinical and Translational Science Awards (CTSA) grant UL1TR004419 from the National Center for Advancing Translational Sciences. Research reported in this publication was also supported by the Office of Research Infrastructure of the National Institutes of Health under award number S10OD030463. The content is solely the responsibility of the authors and does not necessarily represent the official views of the National Institutes of Health. This work was also supported by a grant from the National Human Genome Research Institute (K99HG013547) to J.G.G.

This research has been conducted using the UK Biobank Resource under application number 18177 to P.F.O. Figure 1 was created using the resource BioRender.com.

## Data Availability

The Nextflow pipeline to implement GSR adjustment and perform the analyses in the present study is available at https://github.com/accote45/pathway_overlap.

## Author Contributions Statement

A.C.C. and P.F.O. conceived and designed the study. A.C.C. and R.K. performed the data analysis. J.G.G. processed the gene expression data used in this study and provided input on analysis and interpretation.

A.C.C. wrote the manuscript with inputs from all co-authors. All authors contributed to the interpretation of results and approve the final manuscript.

## Competing Interests Statement

The authors declare no competing interests.

**Supplementary File 1.** Supplementary methods, figures, and tables.

**Supplementary File 2**. All spearman correlations between pathway rankings and external validation evidence.

**Supplementary File 3**. Original and GSR-adjusted MAGMA enrichment results.

**Supplementary File 4**. Original and GSR-adjusted Pascal enrichment results.

**Supplementary File 5**. Original and GSR-adjusted PRSet enrichment results.

**Supplementary File 6**. Original and GSR-adjusted GSA-MiXeR enrichment results.

