## Supplementary File 1 for "Modeling pathway overlap increases accuracy of GWAS gene set enrichment"

**Phenotype ascertainment in the UK Biobank cohort**

- **Alkaline phosphatase:** Alkaline phosphatase levels were extracted from Field ID 30610.
- **Body Mass Index**: Body mass index information was extracted from Field ID 21001.
- **Breast cancer:** Breast cancer was defined by ICD-10 code C50 (malignant neoplasm of breast), by ICD-9 code 174 (malignant neoplasm of female breast), or by cancer code 1002 (self-reported breast cancer). Control participants were female (sex = 0) and had no history of cancer, including no breast cancer diagnosis, other malignancies (ICD-10 C codes), carcinoma in situ (D0), neoplasms of uncertain or unspecified behavior (D37-D48), or self-reported cancer.
- **Coronary Artery Disease:** CAD was defined by fatal or nonfatal myocardial infarction or acute ischemic heart disease identified from hospital records (Field IDs 41270, 41202, 41204, 41203, 41205, and 41271; ICD-10 codes I21–I24 and I25.2, ICD-9 codes 410–412), death records (Field IDs 40001 and 40002), self-reported medical history (Field IDs 6150 and 20002), or coronary revascularization procedures (Field IDs 20004, 41200, and 41272). Controls were defined as participants without any CAD diagnosis.
- **Eosinophil percentage**: Eosinophil percentage levels were extracted from Field ID 30210.
- **HDL cholesterol**: HDL cholesterol levels were extracted from Field ID 30760.
- **Major depressive disorder:** Major depressive disorder (MDD) was defined using UK Biobank Mental Health Questionnaire items collected at instance 0. Participants were classified as cases if they endorsed at least 5 of 11 depression-related items, including three essential items indicating that, during their worst depressive episode, symptoms were present most or all of the time (f20436), occurred almost every day (f20439), and caused more than a little impairment in normal roles (f20440); two core symptoms, prolonged loss of interest in normal activities (f20441) or prolonged feelings of sadness or depression (f20446); and six additional symptoms: difficulty concentrating (f20435), thoughts of death (f20437), tiredness (f20449), worthlessness (f20450), sleep change (f20532), and weight change (f20536). Cases were additionally required to endorse at least one core symptom and all three essential items. Participants with professional diagnoses coded in f20544 as addiction or dependency (2), anxiety/panic attacks (3), or another mental health condition (10) were excluded from case status. Controls were defined as participants who did not meet case criteria and had no record of any professional mental health diagnosis in f20544, no self-reported depression, bipolar/mania, post-natal depression, or schizophrenia in f20002, no prior indication of depression or bipolar disorder in f20126, no hospital ICD-10 mood disorder diagnosis (F30-F39) in either primary (f41202) or secondary (f41204) position, no reported antidepressant medication use at baseline (f20003), and no elevated recent depressive symptom burden, defined as a summed score greater than 4 across nine symptom items (f20507, f20508, f20510, f20511, f20513, f20514, f20517, f20518, f20519).
- **Mean platelet thrombocyte volume:** Mean platelet volume levels were extracted from Field ID 30100.
- **Type 2 diabetes:** Cases were defined as individuals with at least one record of ICD-10 code E11 (Type 2 Diabetes Mellitus) or ICD-9 codes 250 (unspecified diabetes), 250.0 (diabetes without complications), or 250.2 (diabetes with renal manifestations). Diagnostic codes were extracted from main diagnoses (data fields 41202 and 41203 for ICD-10 and ICD-9, respectively), secondary diagnoses (fields 41204 and 41205), general diagnoses (fields 41270 and 41271), and cause of death records including both underlying and contributory causes (fields 40001 and 40002). Individuals were excluded if they had any record of Type 1 Diabetes (ICD-10 E10), gestational diabetes (ICD-10 O24; ICD-9 250.1, 250.3, or 648), or diagnosis before age 35 years to capture adult-onset diabetes characteristic of T2D.

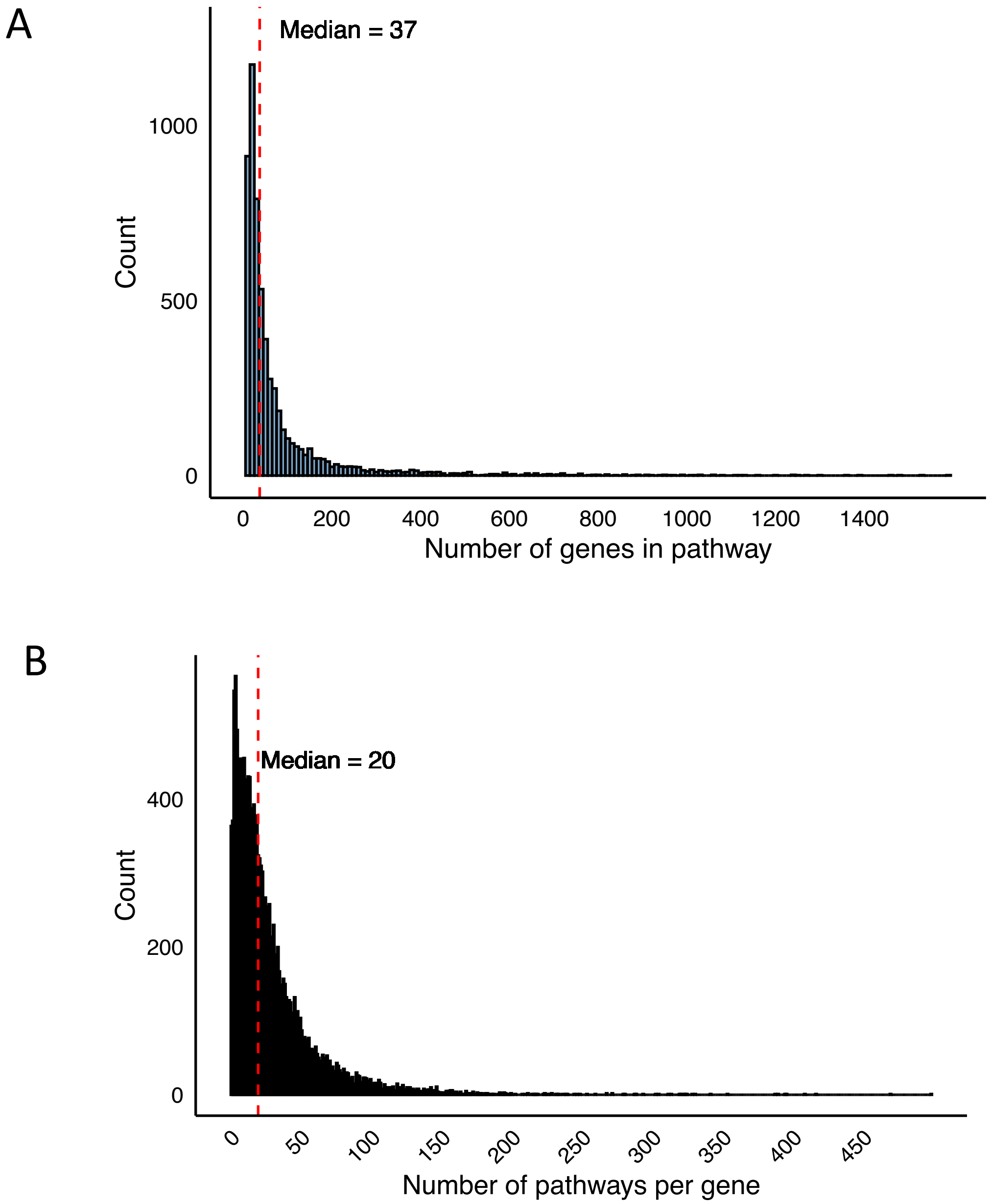


Fig S1. Distribution of A) pathway size and B) gene frequency in the MSigDB database.

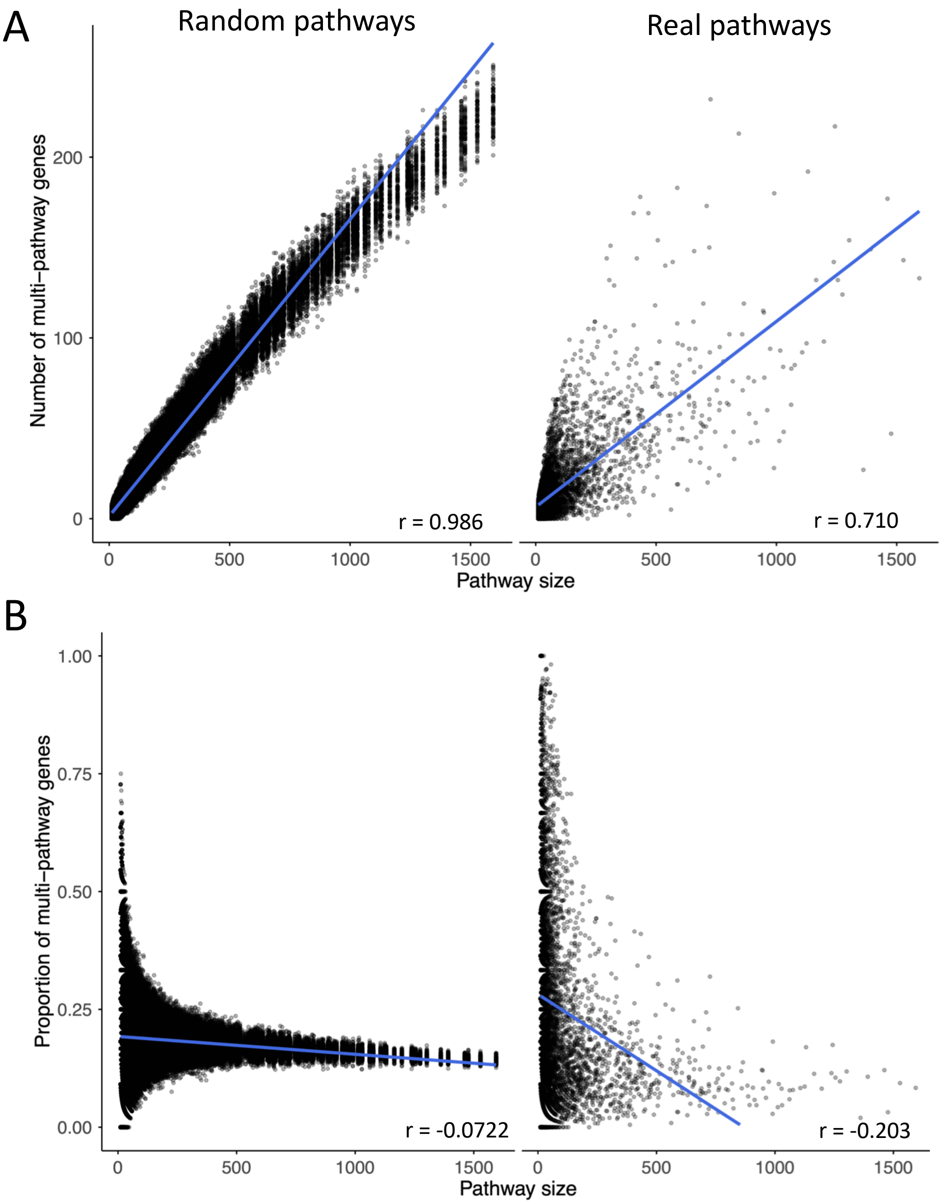


Figure S2. Relationship between pathway size and multi-pathway gene content in both random and real pathway databases. A) Number of multi-pathway genes per pathway. B) Proportion of multi-pathway genes per pathway. Multi-pathway genes were defined as genes present in at least 100 pathways. Multi-pathway proportion is defined as the number of multi-pathway genes divided by the total number of genes in each pathway. Randomized pathways were generated via GSR and downsampled to the first 100 permutations for visualization.

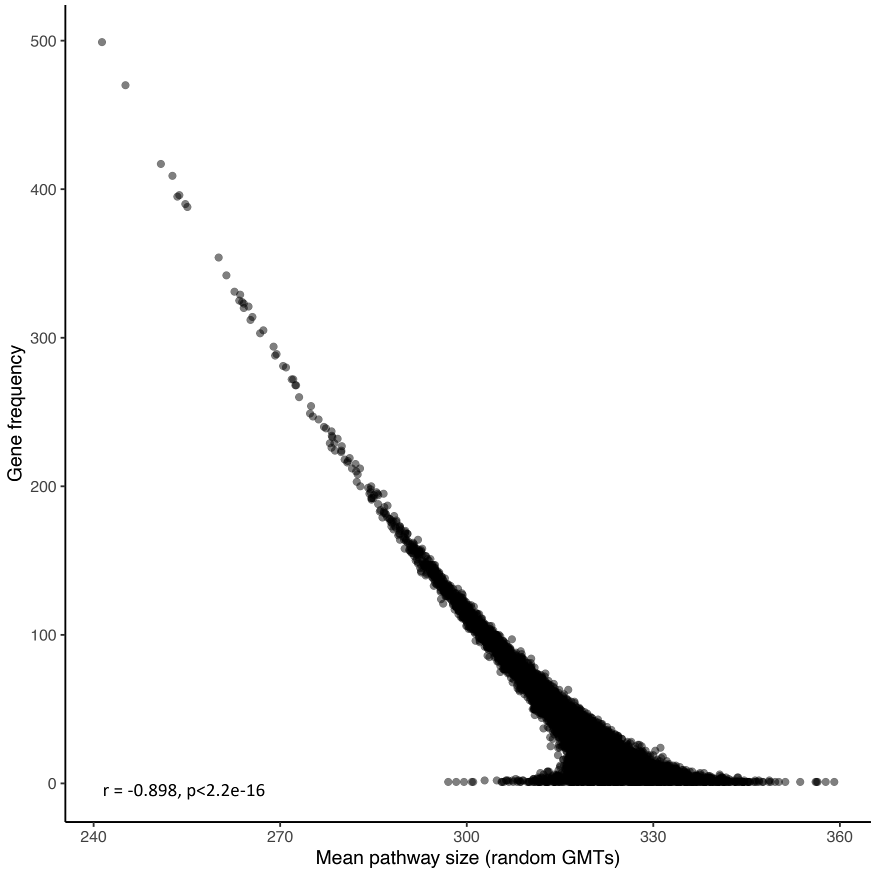


Fig S3. Association between a gene’s frequency and the mean size of random pathways containing that gene.

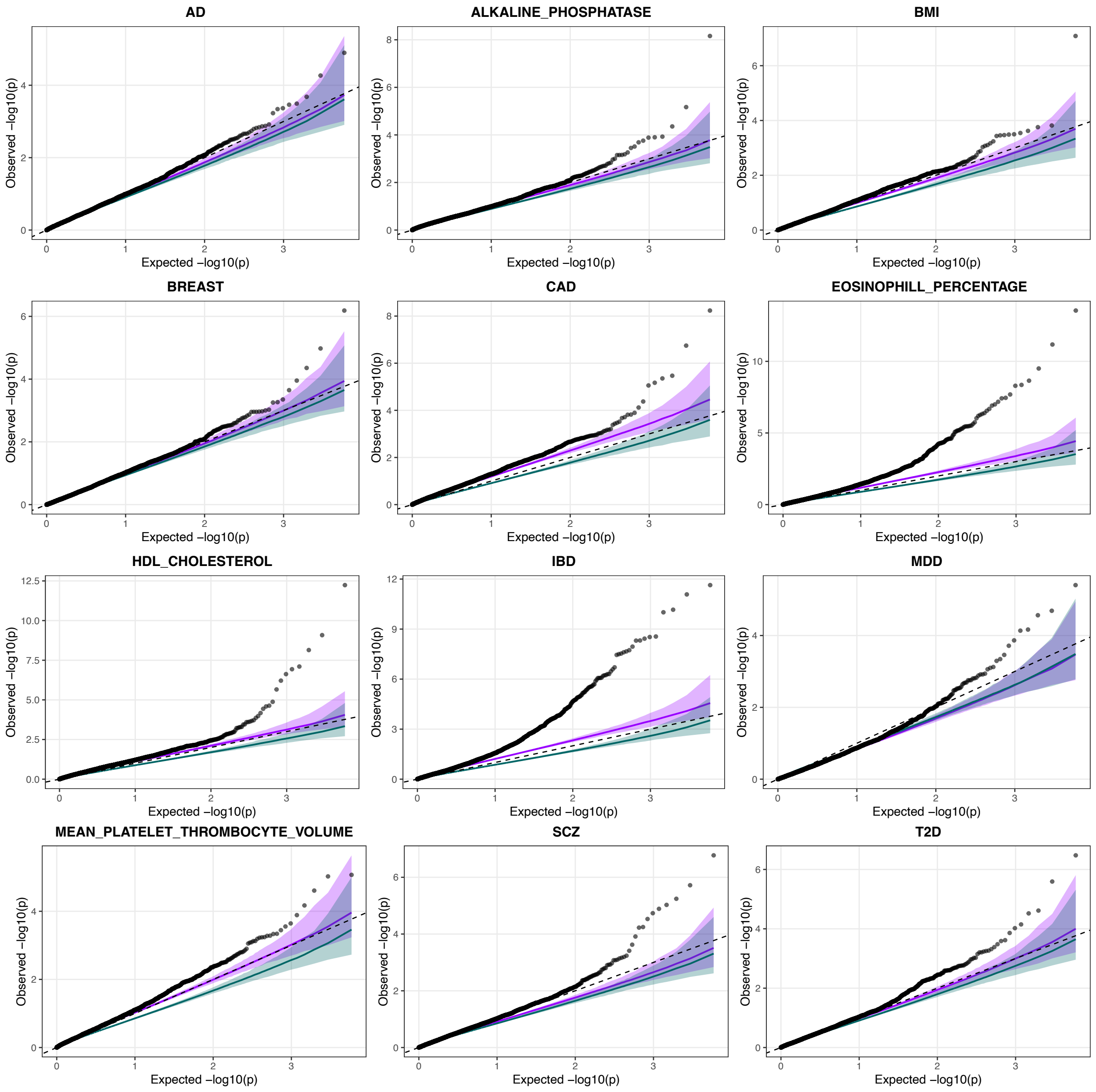


Fig S4. QQ plots comparing observed MAGMA pathway enrichment p-values (black points) to two null models: GSR (purple) and PS (green), across multiple traits. Lines show median –log10(p-value) at a given rank position, and shaded regions show 2.5%-97.5% percentile bands at a given rank position (95% of the 1000 permuted p-values at the same rank fall within the shaded region). The dashed line indicates expectation under a uniform null distribution.

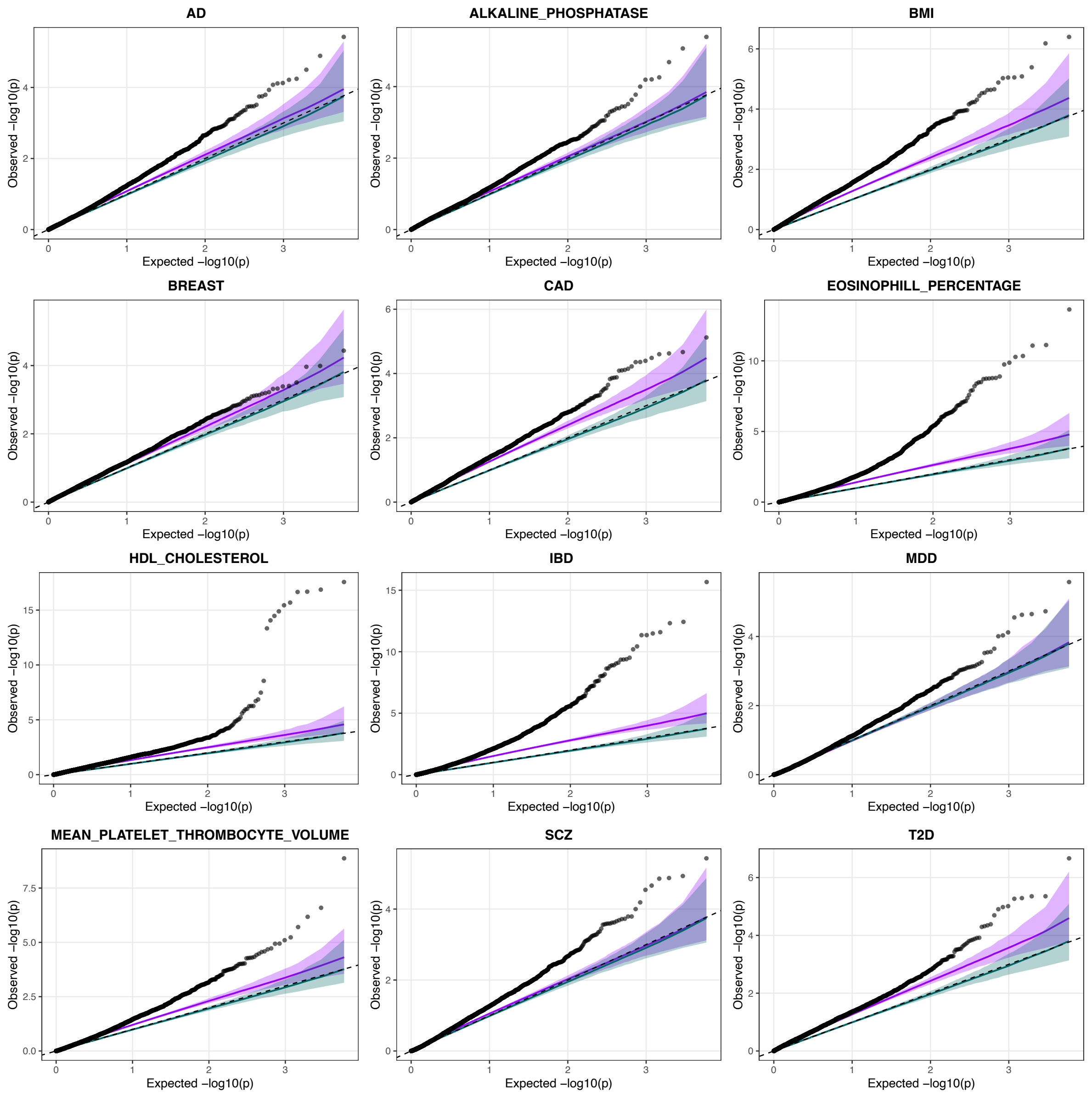


Fig S5. QQ plots comparing observed Pascal pathway enrichment p-values (black points) to two null models: GSR (purple) and PS (green), across multiple traits. Lines show median –log10(p-value) at a given rank position, and shaded regions show 2.5%-97.5% percentile bands at a given rank position (95% of the 1000 permuted p-values at the same rank fall within the shaded region). The dashed line indicates expectation under a uniform null distribution.

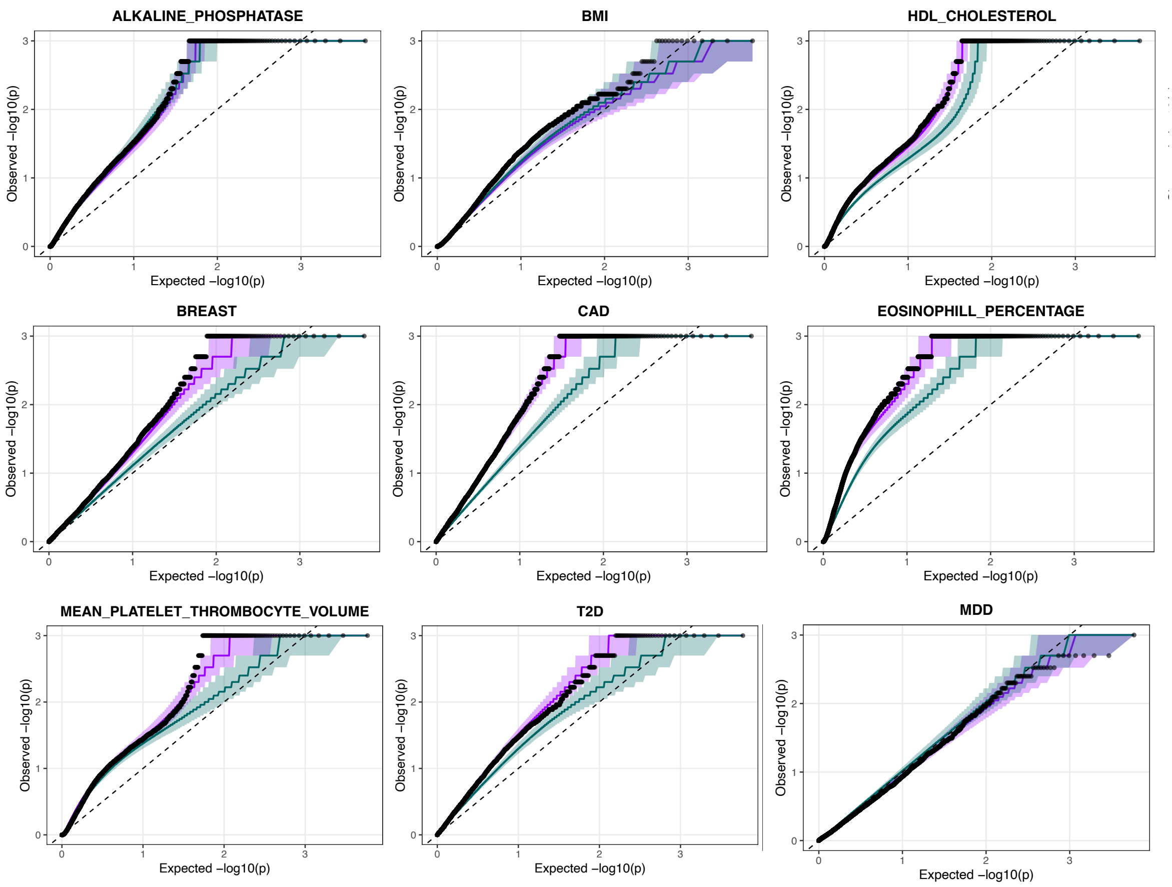


Fig S6. QQ plots comparing observed PRSet pathway enrichment p-values (black points) to two null models: GSR (purple) and PS (green), across multiple traits. The observed p-values shown are the competitive p-values output from the PRSet tool after internal permutation testing to adjust for pathway size. Lines show median –log10(p-value) at a given rank position, and shaded regions show 2.5%-97.5% percentile bands at a given rank position (95% of the 1000 permuted p-values at the same rank fall within the shaded region). The dashed line indicates expectation under a uniform null distribution.

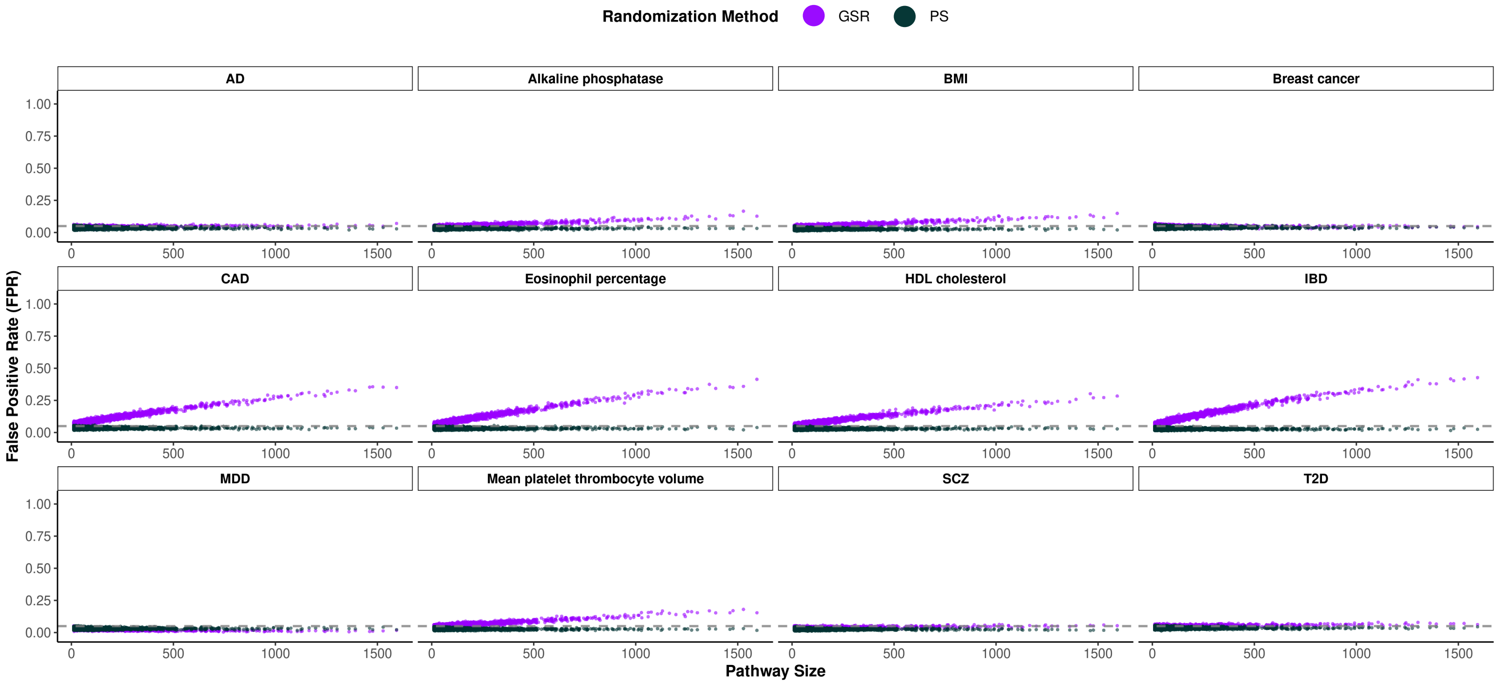


Fig S7. For MAGMA enrichment analysis, the relationship between pathway size and FPR as estimated from random pathways for all GWAS traits. PS = Null model preserving pathway size, GSR = Gene swap randomization null model preserving pathway size and gene frequency.

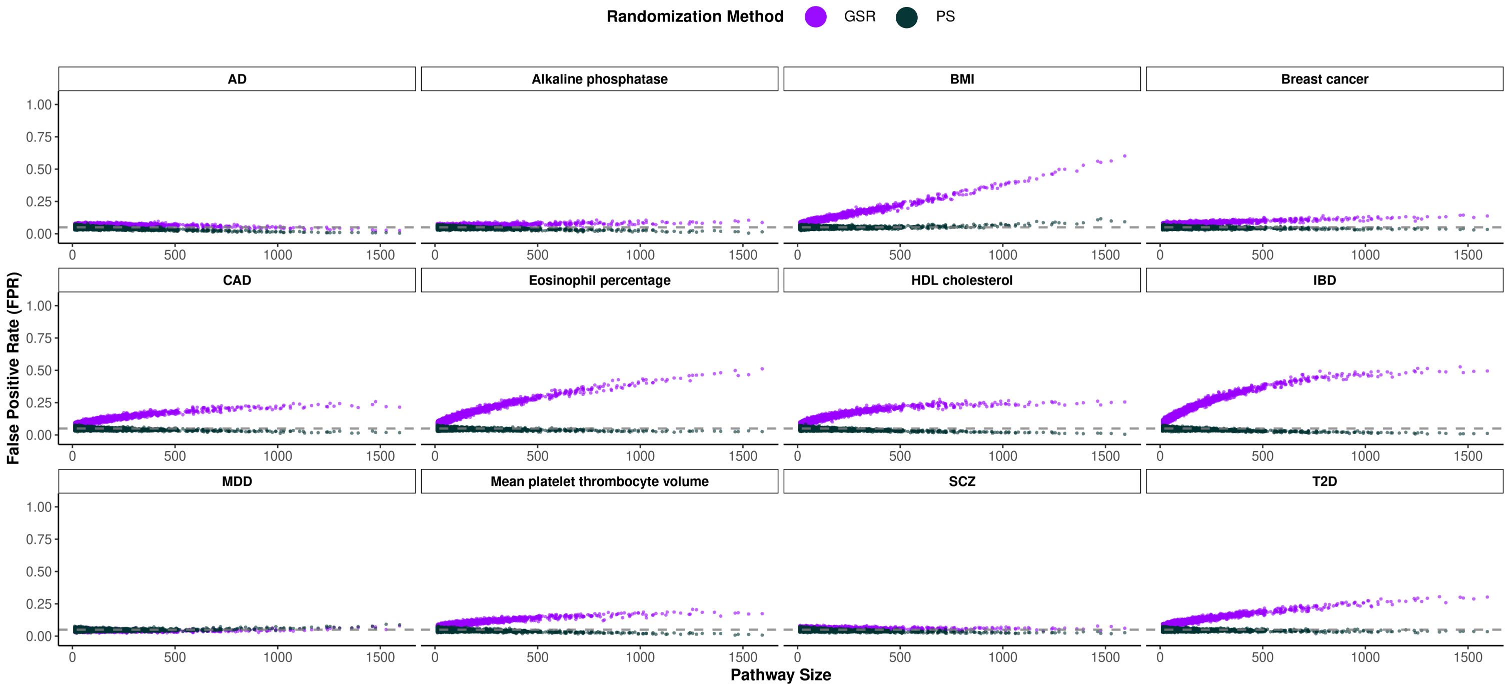


Fig S8. For Pascal enrichment analysis, the relationship between pathway size and FPR as estimated from random pathways for all GWAS traits. PS = Null model preserving pathway size, GSR = Gene swap randomization null model preserving pathway size and gene frequency.

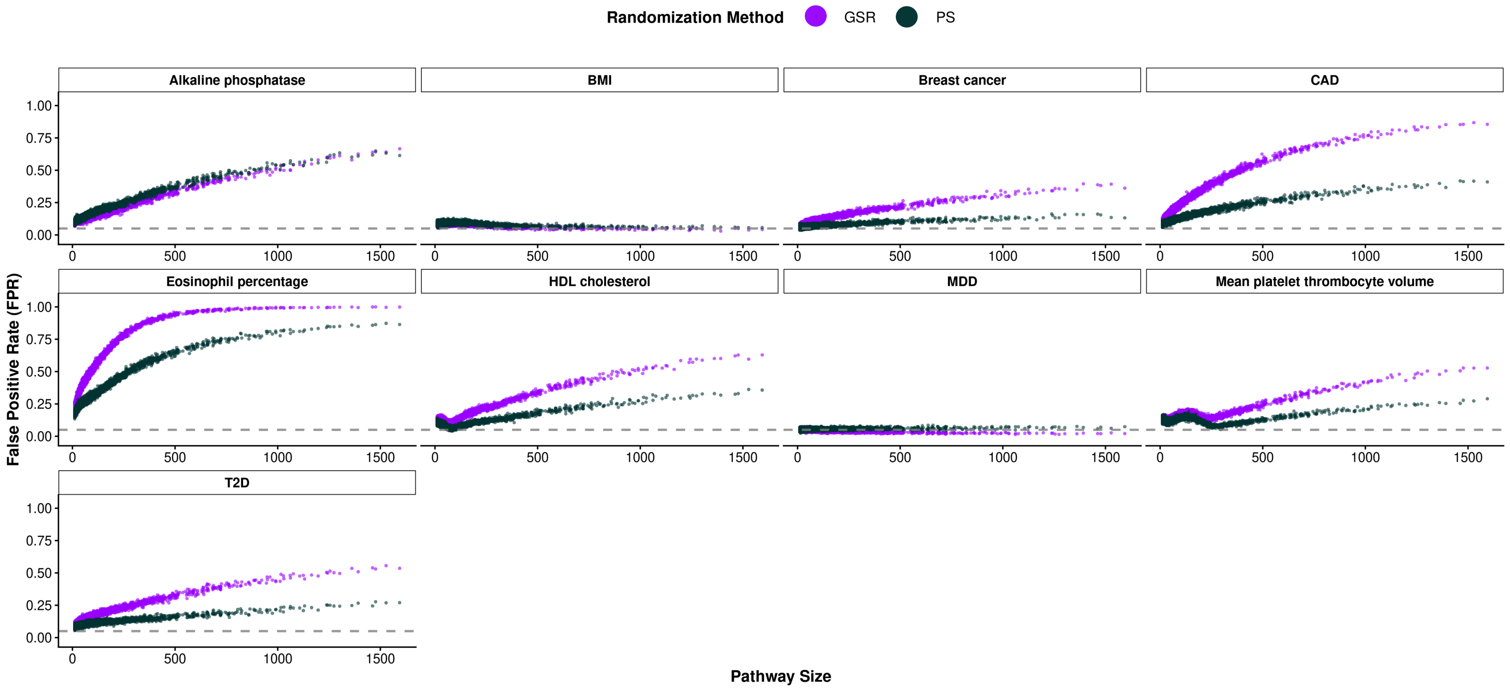


Fig S9. For PRSet analysis, the relationship between pathway size and FPR as estimated from random pathways for all GWAS traits. PS = Null model preserving pathway size, GSR = Gene swap randomization null model preserving pathway size and gene frequency.

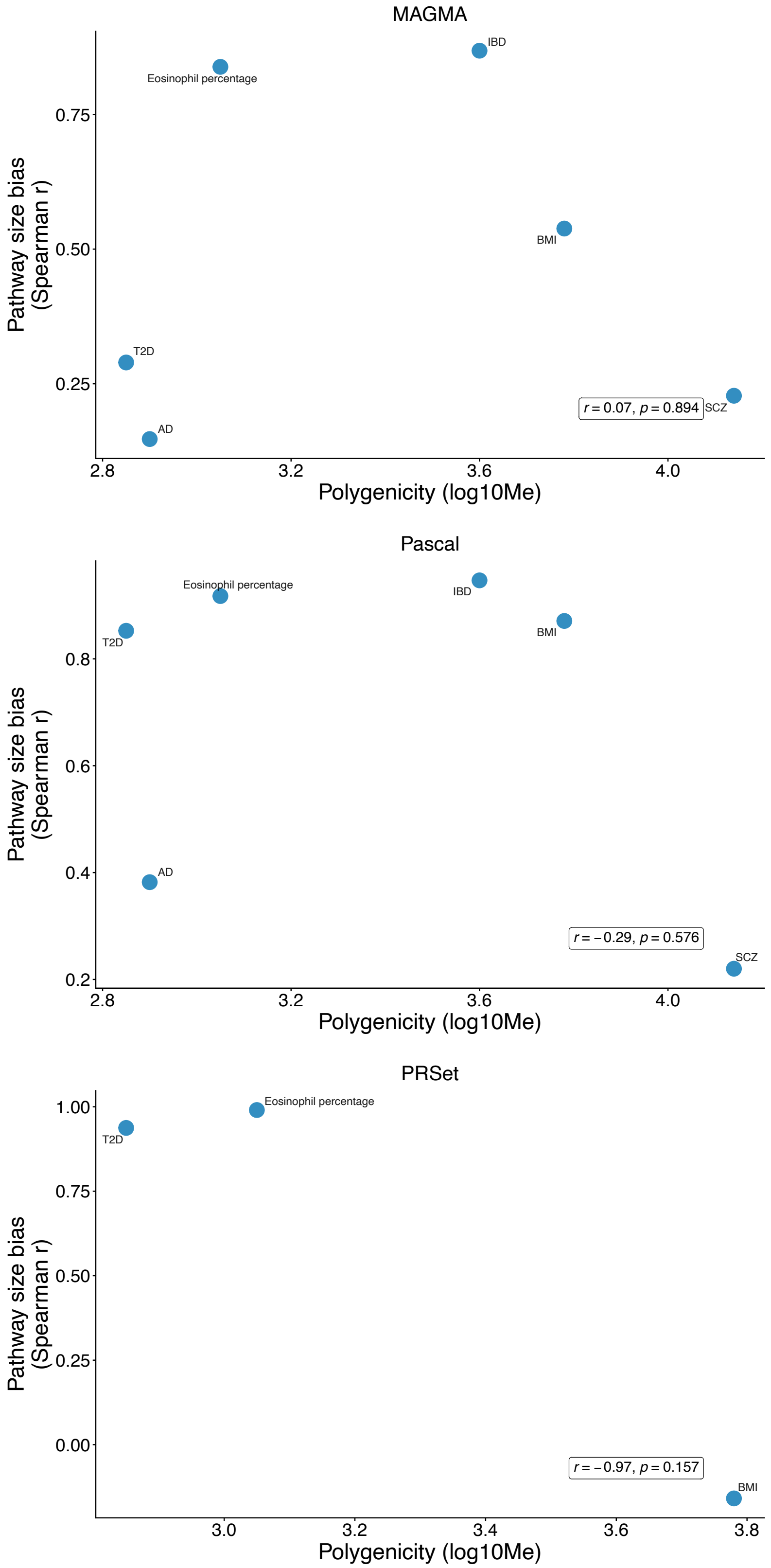


Figure S10. Relationship between polygenicity and pathway size bias across gene-set analysis methods. Scatterplots showing the correlation between polygenicity ($log_{10}$Me) and pathway size bias (measured as Spearman correlation between pathway size and false positive rate) for MAGMA, Pascal, and PRSet. Pearson correlation coefficients and p-values are displayed in the bottom right of each panel. Polygenicity estimates were taken from O'Connor et al. (*AJHG*, 2019; doi: 10.1016/j.ajhg.2019.07.003).

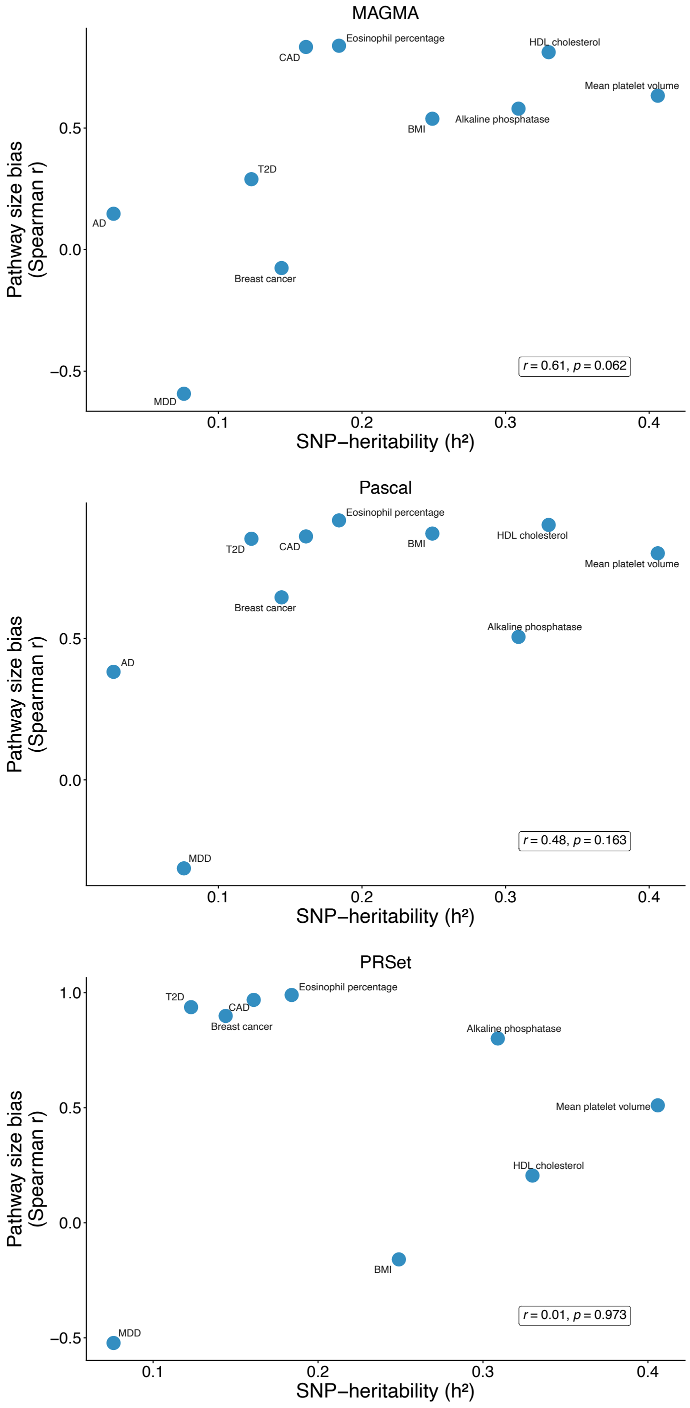


Figure S11. Relationship between SNP-heritability and pathway size bias across gene-set analysis methods. Scatterplots showing the correlation between SNP-heritability ($h^2$) and pathway size bias (measured as Spearman correlation between pathway size and false positive rate) for MAGMA, Pascal, and PRSet. Pearson correlation coefficients and p-values are displayed in the bottom right of each panel. Traits with non-significant heritability (IBD, SCZ h2 $p<0.05$) were excluded. SNP heritability values taken from UKB estimates from the Neale Lab data browser.

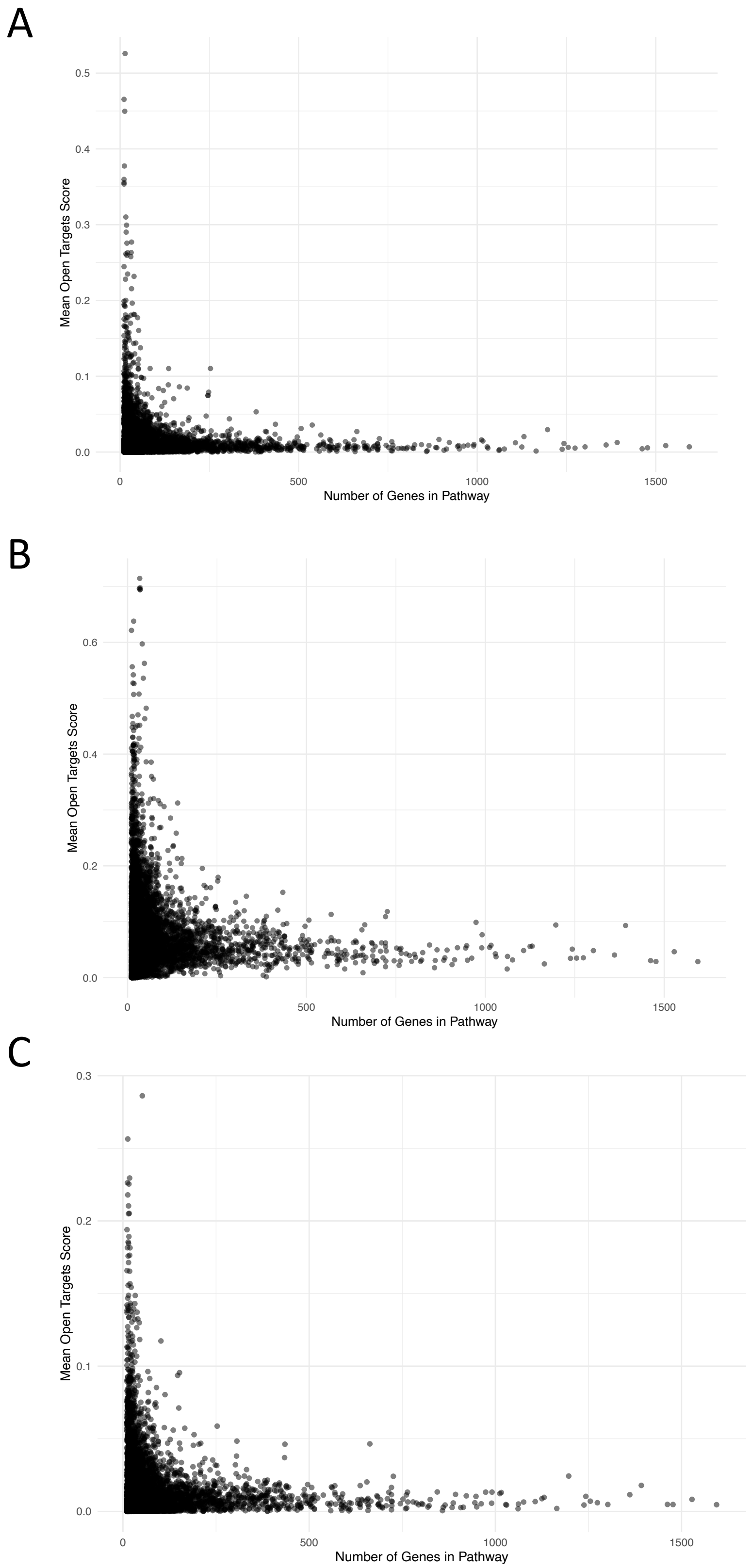


Fig S12. Relationship between pathway size and mean Open Targets score for three representative traits: A) Alzheimer's disease, B) coronary artery disease, and C) inflammatory bowel disease.

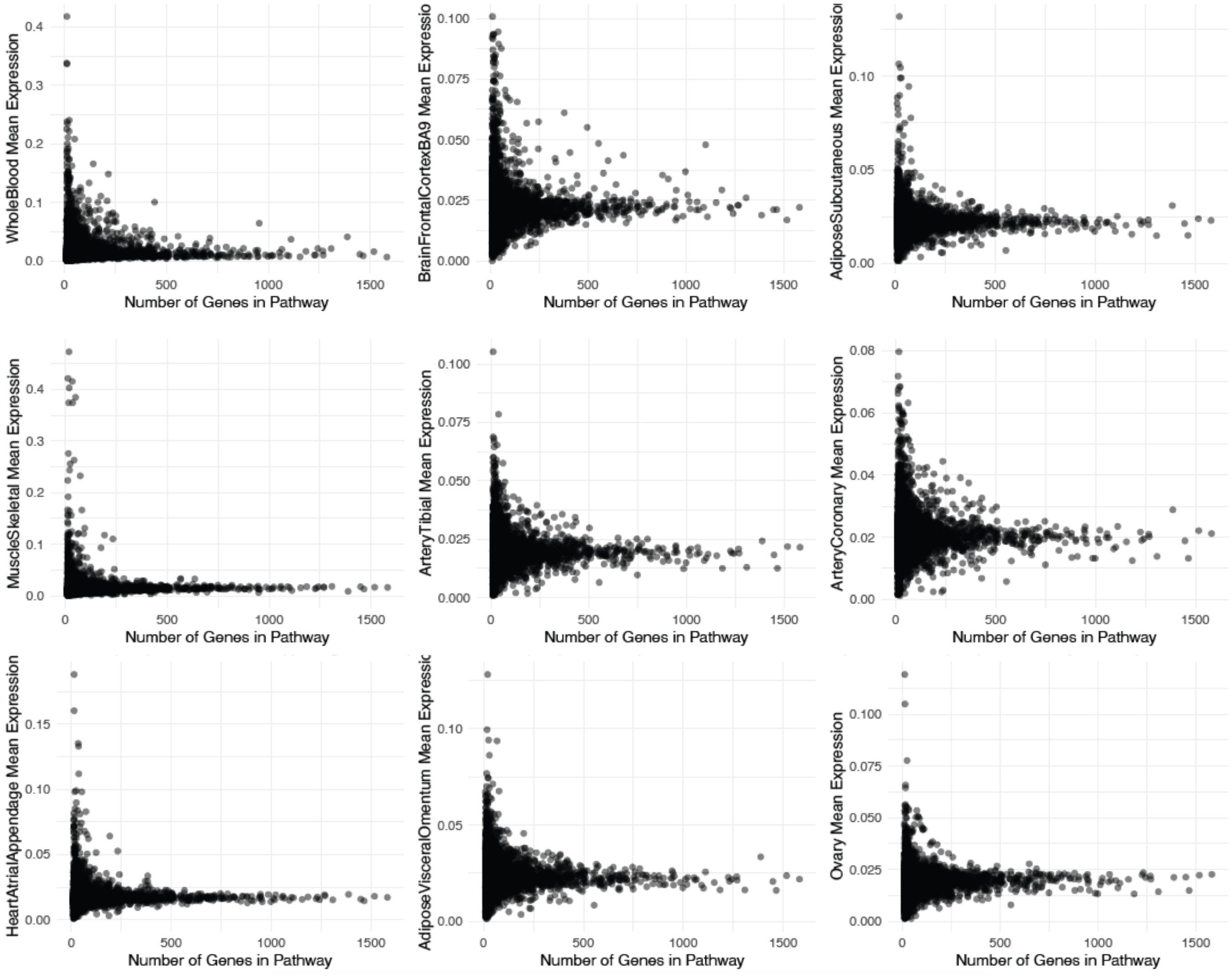


Fig S13. Relationship between pathway size and mean tissue specificity score for nine example GTEx tissues. Mean expression = Mean tissue expression specificity score per pathway.

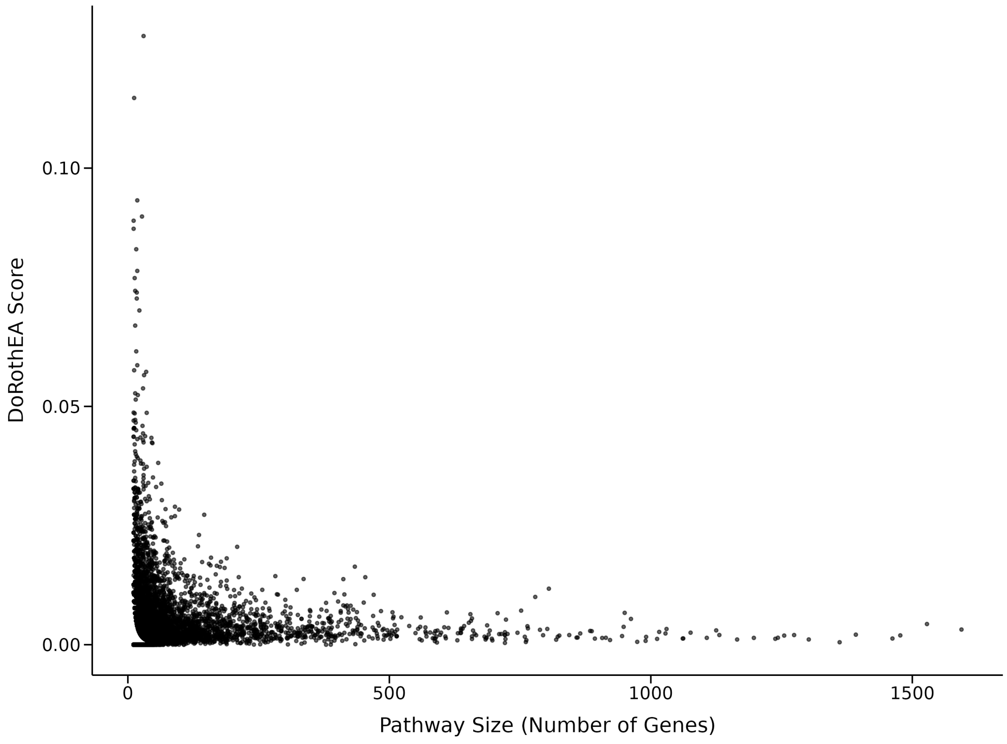


Fig S14. Relationship between pathway size and mean DoRothEA score.

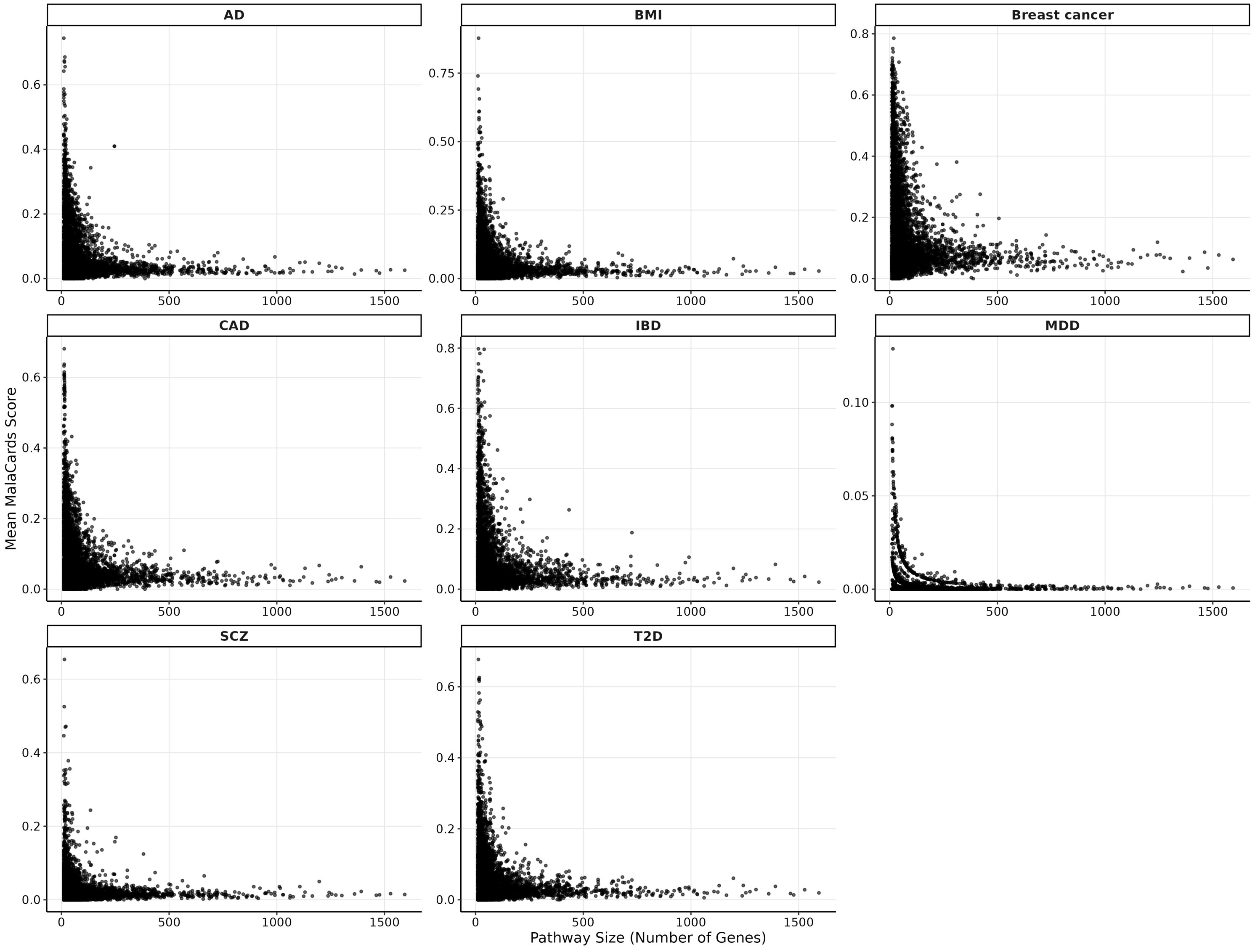


Fig S15. Relationship between pathway size and mean Malacards disease association score.

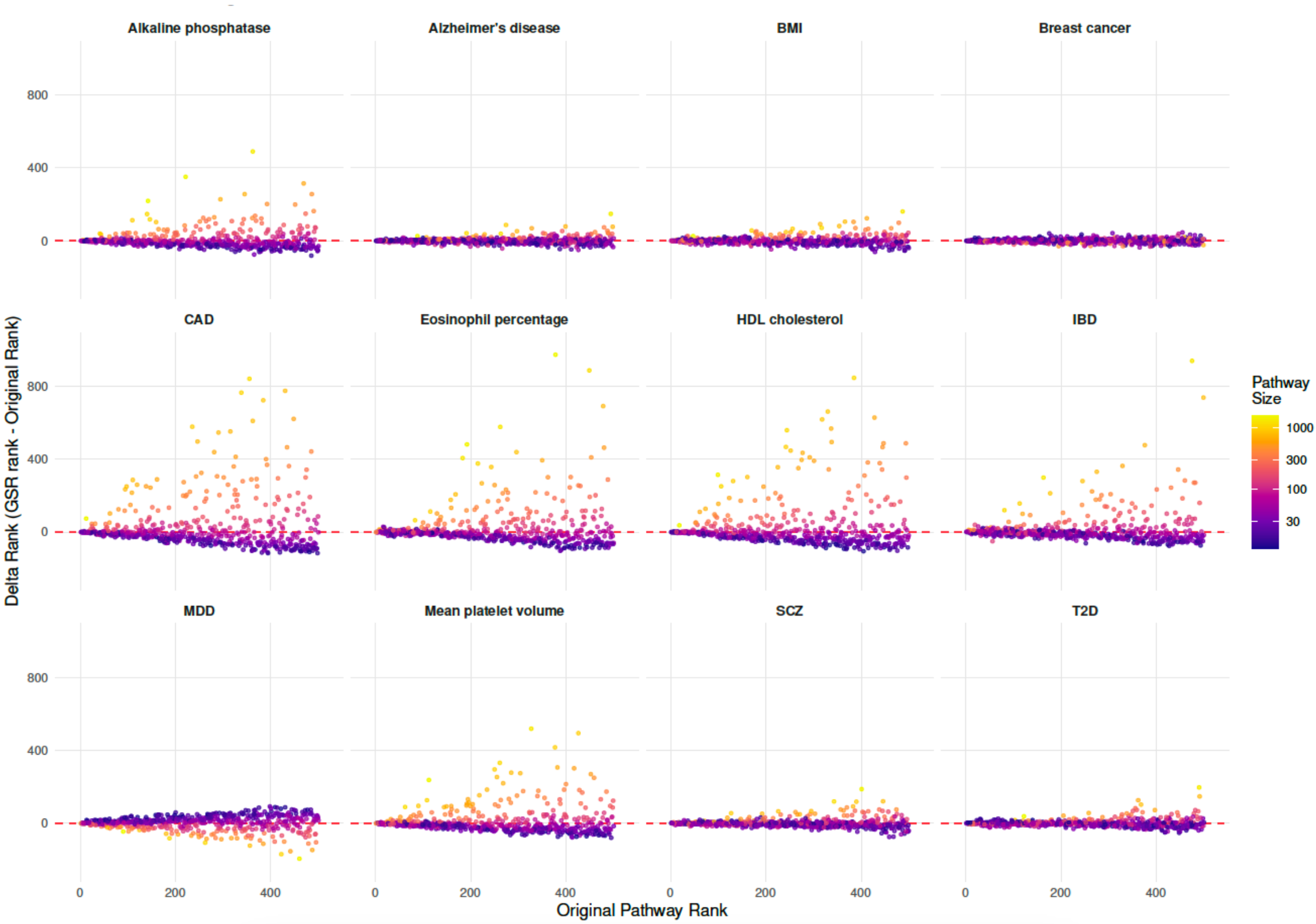


Fig S16. For the top 500 pathways ranked by MAGMA significance for each trait, each panel shows the relationship between a pathway's original MAGMA rank and its change in rank after calibration (Δ rank = GSR rank – original rank). Each point represents a pathway and is colored by pathway size (number of genes). Positive Δ rank values indicate a pathway was downweighted after calibration, whereas negative values indicate upward re-ranking. The dashed red line denotes no change in rank (Δ rank = 0).

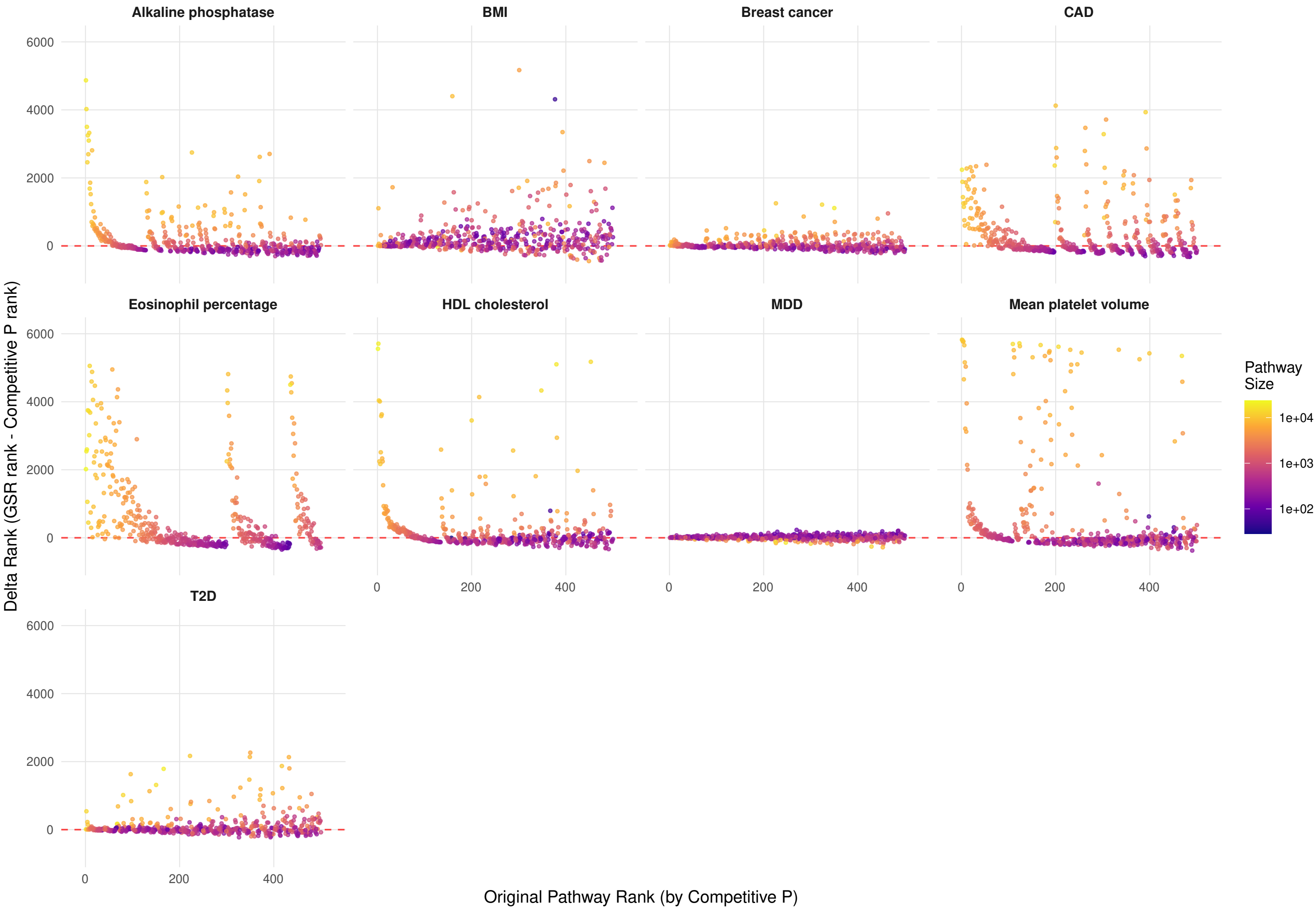


Fig S17. For the top 500 pathways ranked by PRSet significance for each trait, each panel shows the relationship between a pathway's original PRSet rank and its change in rank after calibration (Δ rank = GSR rank – original rank). Original PRSet rank was defined by the competitive p-value output by the PRSet tool. Each point represents a pathway and is colored by pathway size (number of independent SNPs). Positive Δ rank values indicate a pathway was downweighted after calibration, whereas negative values indicate upward re-ranking. The dashed red line denotes no change in rank (Δ rank = 0).

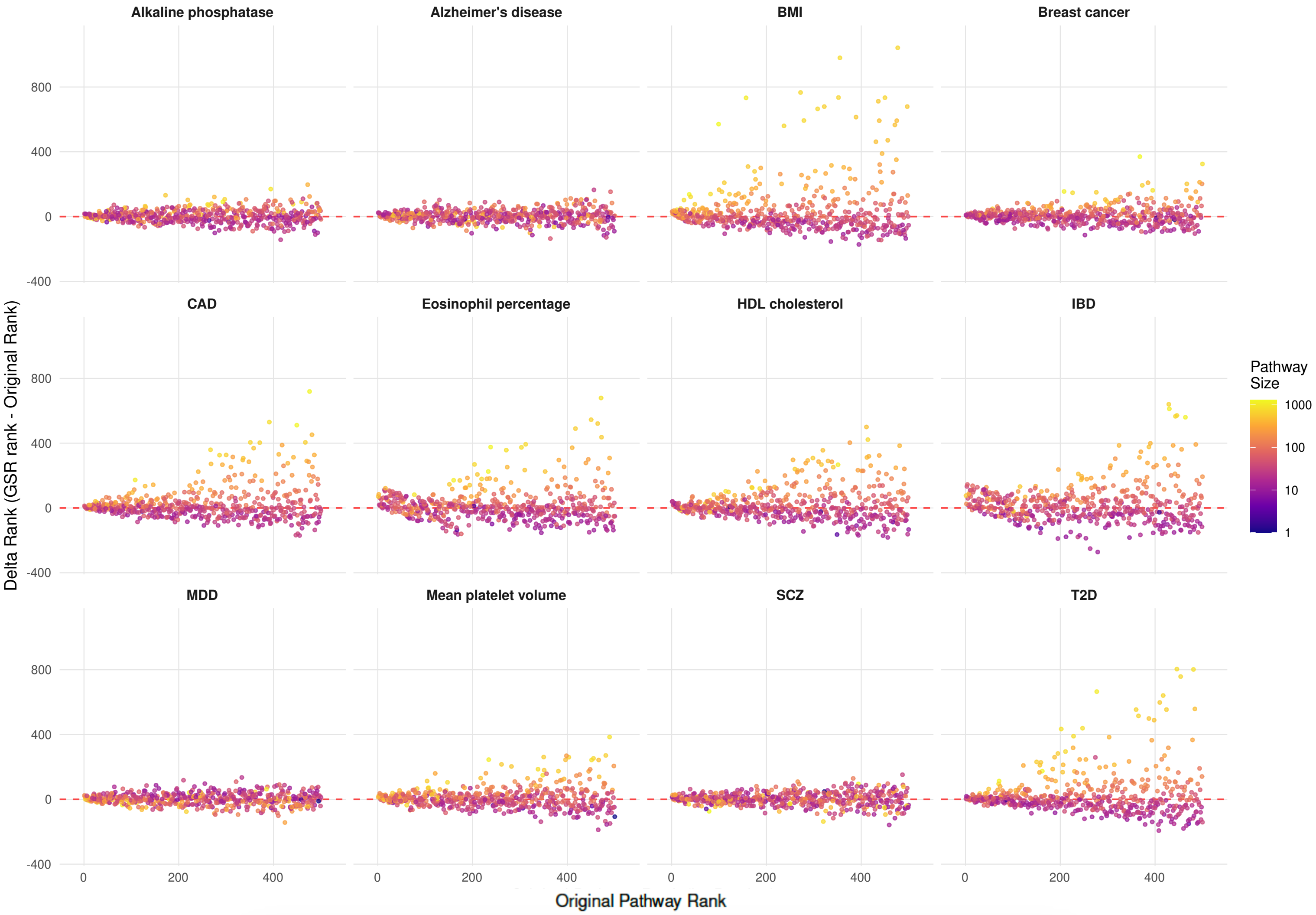


Fig S18. For the top 500 pathways ranked by Pascal significance for each trait, each panel shows the relationship between a pathway's original Pascal rank and its change in rank after calibration (Δ rank = GSR rank – original rank). Each point represents a pathway and is colored by pathway size (number of genes). Positive Δ rank values indicate a pathway was downweighted after calibration, whereas negative values indicate upward re-ranking. The dashed red line denotes no change in rank (Δ rank = 0).

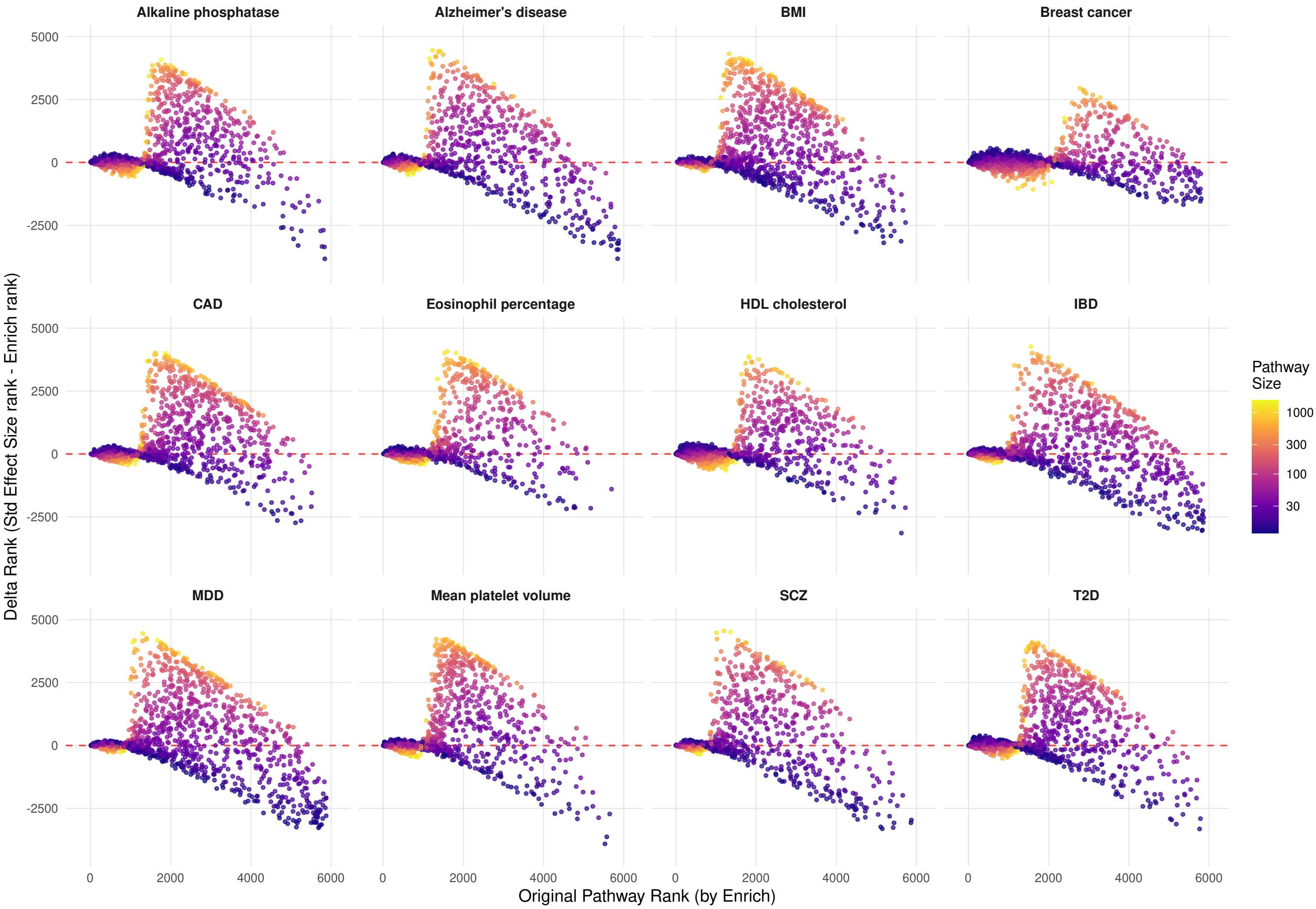


Fig S19. Each panel shows the relationship between a pathway's original GSA-MiXeR rank and its change in rank after calibration (Δ rank = GSR rank – original GSA-MiXeR rank). Each point represents a pathway and is colored by pathway size (number of genes). Positive Δ rank values indicate a pathway was downweighted after calibration, whereas negative values indicate upward re-ranking. The dashed red line denotes no change in rank (Δ rank = 0).

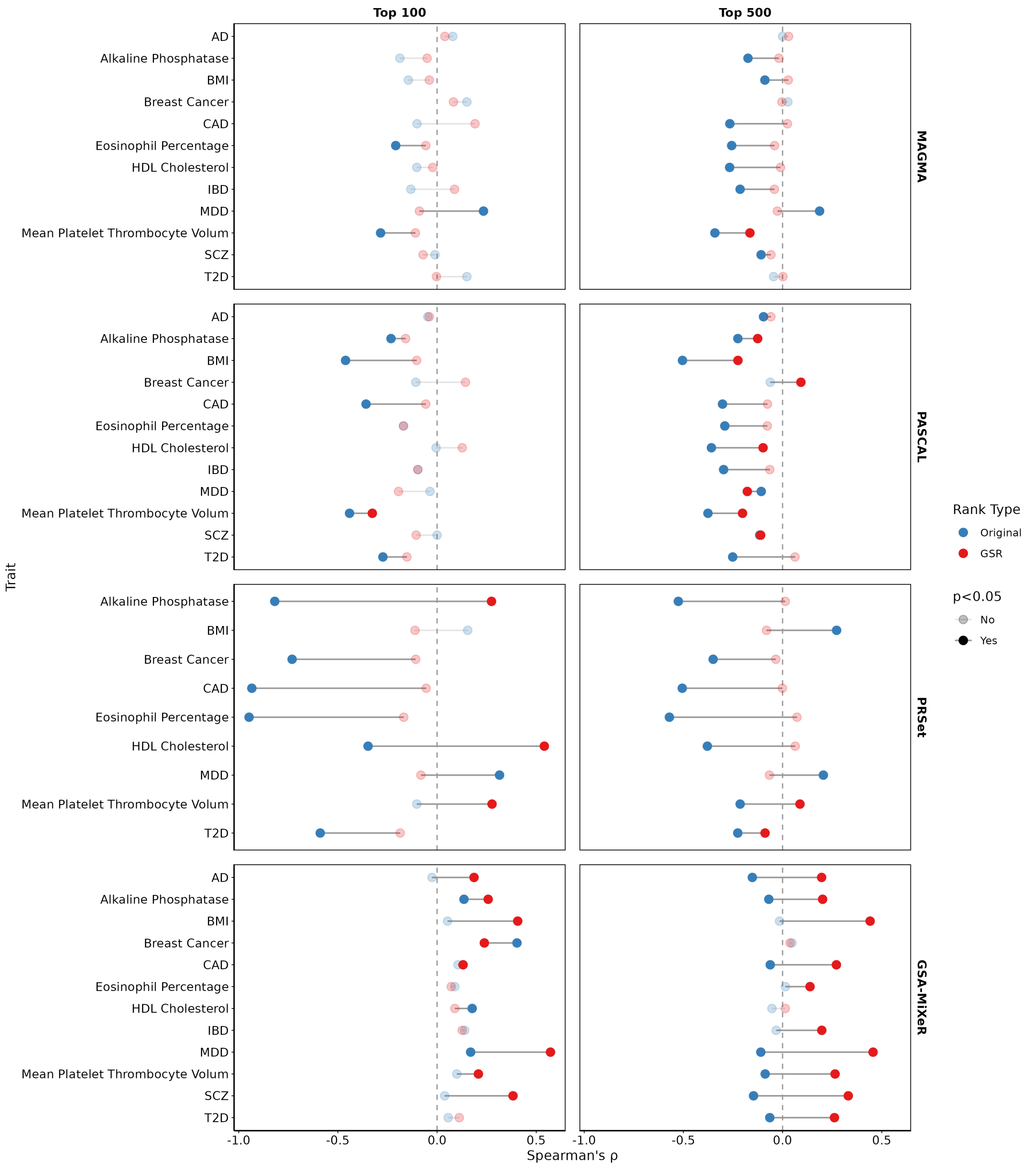


Fig S20. Spearman rank correlation between pathway size and pathway enrichment rank, calculated for the top 100 and top 500 pathways across the four enrichment tools. Blue points show correlation with the original, unadjusted enrichment ranking, and red points show correlation after GSR adjustment. Negative ρ values indicated a pathway size bias where larger gene sets tend to show more significant enrichment. Filled points denote nominally significant correlation ($p<0.05$).

Table S1. GWAS summary statistics used in gene set enrichment analyses

| Source | GWAS Trait | Category | N (case/control) |
|---|---|---|---|
| Verma 2024 | Body mass index | anthropometric, cardiometabolic | 424,221 |
| Nikpay 2015 | Coronary artery disease | cardiometabolic | 184,305 (60,801/123,504) |
| Scott 2017 | Type 2 diabetes | cardiometabolic | 159,208 (26,676/132,532) |
| Adams 2025 | Major depressive disorder | psychiatric | 1,639,572 (357,636/1,281,936) |
| Wightman 2021 | Alzheimer's disease | neurodegenerative | 398,058 (39,918/358,140) |
| Trubetskoy 2022 | Schizophrenia | psychiatric | 132,644 (53,386/77,258) |
| de Lange 2017 | Inflammatory bowel disease | immune | 59957 (25,042/34,915) |
| Zhang 2020 | Breast cancer | cancer | 247,173 (133,384/113,789) |
| UKB | HDL cholesterol | lab value | 173,200 |
| UKB | Mean platelet thrombocyte volume | lab value | 173,200 |
| UKB | Alkaline phosphatase | lab value | 173,200 |
| UKB | Eosinophil percentage | lab value | 173,200 |

Table S2. UK Biobank cohorts used as target samples in PRSet analyses

| GWAS Trait | N (case/control) |
|---|---|
| BMI | 93,511 |
| CAD | 93,511 (5,307/88,204) |
| T2D | 93, 291 (4,994/88,297) |
| MDD | 61,635 (8,207/53,428) |
| Breast cancer | 42,811 (2,475/40,336) |
| HDL cholesterol | 93,511 |
| Mean platelet thrombocyte volume | 93,511 |
| Alkaline phosphatase | 93,511 |
| Eosinophil percentage | 93,511 |

Table S3. Phenotypes used in Open Targets correlation analysis

| GWAS trait | Disease Ontology ID | N pathway genes with Open Targets evidence |
|---|---|---|
| IBD | EFO_0000555 | 594 |
| Breast cancer | MONDO_0007254 | 7164 |
| AD | MONDO_0004975 | 534 |
| SCZ | MONDO_0005090 | 3944 |
| MDD | MONDO_0002050 | 267 |
| T2D | MONDO_0005148 | 5761 |
| CAD | EFO_0001645 | 2601 |
| BMI | EFO_0001073 | 400 |

Table S4. Phenotypes used in Malacards correlation analysis

| GWAS trait | MCID | N pathway genes with Malacards evidence | Name in Malacards database |
|---|---|---|---|
| AD | ALZ001 | 673 | Alzheimer's disease |
| BMI | BDY004 | 891 | Body Mass Index Quantitative Trait Locus 11 |
| Breast cancer | BRS047 | 1899 | Breast Cancer |
| CAD | HRT032 | 806 | Heart disease |
| | CRN162 | 11 | Coronary Atherosclerosis |
| | CRN018 | 318 | Coronary Artery Anomaly |
| | MYC007 | 400 | Myocardial infarction |
| IBD | INF037 | 1110 | Inflammatory Bowel Disease |
| MDD | MJR001 | 35 | Major Depressive Disorder |
| | DPR007 | 104 | Depressive disorder |
| SCZ | SCH015 | 578 | Schizophrenia |
| T2D | TYP009 | 689 | Type 2 Diabetes Mellitus |

Table S5. GTEx tissues tested for each GWAS trait in the tissue-specificity correlation analysis

| GWAS trait | GTEx tissue |
|---|---|
| AD | Hippocampus |
| Alkaline phosphatase | Liver |
| BMI | Brain Frontal Cortex BA9 |
| Breast cancer | Breast Mammary Tissue |
| CAD | Artery Coronary |
| Eosinophil percentage | Whole blood |
| HDL cholesterol | Liver |
| IBD | Small intestine terminal ileum |
| MDD | Brain Frontal Cortex BA9 |
| Mean platelet thrombocyte volume | Whole blood |
| Schizophrenia | Brain Frontal cortex BA9 |
| T2D | Pancreas |

Table S5. SNP heritability and polygenicity estimates for GWAS traits

| GWAS trait | SNP heritability* | | | Common variant polygenicity** | |
|---|---|---|---|---|---|
| | Phenotype | h2 | h2 pvalue | Phenotype | log10Me Common |
| AD | Illnesses of mother: Alzheimer's disease/dementia | 0.027 | 0.000737 | Alzheimer's | 2.900 |
| Alkaline phosphatase | Alkaline phosphatase (U/L) | 0.309 | 0.00000202 | | |
| BMI | Body mass index (BMI) | 0.249 | 2.52E-194 | BMI | 3.780 |
| Breast cancer | Cancer code, self-reported: breast cancer | 0.144 | 8.15E-09 | | |
| CAD | coronary atherosclerosis | 0.161 | 1.98E-15 | | |
| Eosinophil percentage | Eosinophill count | 0.184 | 4.85E-44 | Eosinophil count | 3.050 |
| HDL cholesterol | HDL cholesterol (mmol/L) | 0.330 | 0.0000017 | | |
| IBD | Non-cancer illness code, self-reported: inflammatory bowel disease | 0.161 | 0.335 | IBD | 3.600 |
| MDD | non-cancer illness code, self-reported: depression | 0.076 | 1.59E-13 | | |
| Mean platelet thrombocyte volume | Mean platelet (thrombocyte) volume | 0.406 | 1.11E-12 | | |
| SCZ | Mental health problems ever diagnosed by a professional: Schizophrenia | 0.777 | 0.108 | Schizophrenia | 4.140 |
| T2D | Non-cancer illness code, self-reported: type 2 diabetes | 0.123 | 0.00274 | Type II diabetes | 2.850 |

*SNP heritability estimates taken from Neale lab UKB Heritability portal (nealelab.github.io/UKBB_ldsc/)
**Polygenicity estimates taken from O'Connor et al. (*AJHG*, 2019; doi: 10.1016/j.ajhg.2019.07.003)
